# Disorder-specific genetic effects drive the associations between psychopathology and cognitive functioning

**DOI:** 10.1101/2025.06.06.25329135

**Authors:** Wangjingyi Liao, Engin Keser, Andrea Allegrini, Kaili Rimfeld, Robert Plomin, Margherita Malanchini

## Abstract

**Background:** Cognitive functioning is a critical dimension of psychopathology that remains under-investigated. Because cognitive deficits often transcend diagnostic boundaries, it has been challenging to delineate specific relationships between psychiatric disorders and cognitive functioning. Genetic research offers novel, powerful tools to disentangle shared (transdiagnostic) from disorder-specific effects, opening new avenues for understanding how psychopathology relates to cognitive functioning.

**Method:** We used genomic structural equation modelling and derived polygenic scores in the Twins Early Development Study sample (N = 7764) to examine associations between genetic risk for psychopathology and cognitive functioning across development. We explored the extent to which differences in cognitive skills (general cognitive ability, verbal and non-verbal abilities) are associated with transdiagnostic versus disorder-specific risk.

**Results:** The results showed that the relationship between psychopathology and cognitive functioning is primarily driven by disorder-specific genetic effects, rather than by transdiagnostic factors. While for some psychiatric disorders (e.g., ADHD, Tourette’s) associations with cognitive skills were negative, for others (e.g., autism spectrum disorder, anorexia nervosa) they were positive. Associations increased across development and varied for different cognitive skills. Within-siblings analyses indicated that genetic associations operated through distinct pathways: for some disorders (e.g., ADHD) effects were largely shared between siblings while for others (e.g., ASD) they were specific to each individual within a family.

**Conclusions:** In contrast to psychiatric symptoms, which are most effectively predicted by transdiagnostic genetic risk for psychopathology, our findings highlight the need to consider disorder-specific genetic effects when examining associations with cognitive functioning. Research and interventions focused solely on transdiagnostic risk are unlikely to capture the full complexity of these relationships or to enhance our understanding of the distinct cognitive profiles associated with different psychiatric conditions.

## Introduction

Cognitive development is characterized by age-related increases in several cognitive functions, including vocabulary, complex reasoning, and learning. Variation in cognitive development is associated with key societal and medical outcomes throughout the lifespan, including physical and mental health (1, 2). Studies have highlighted the positive association between cognitive functioning and mental health outcomes across the life span (3): higher cognitive competencies have been consistently associated with better mental health (4). An association between cognition and mental health is also observed at the other end of the spectrum of health and ability, as cognitive deficits are common in patients diagnosed with psychiatric disorders (5).

Despite this well-established association between cognition and mental health, cognitive functioning remains an under-investigated dimension of psychopathology, if compared to affective and emotional symptoms (6). Cognitive deficits often transcend traditional diagnostic boundaries, in the same way that core affective and emotional symptoms are shared across multiple conditions (7). For example, an inability to forget or inhibit thoughts, is central to both post-traumatic stress disorder (PTSD) and obsessive-compulsive disorder (OCD), where patients experience intrusive thoughts or intense flashbacks. Cognitive deficits in attention, memory, and language are also observed across schizophrenia, bipolar disorder, and autism spectrum disorder (ASD; see Millan et al., 2012 (6) for a review).

This overlap between cognitive symptoms across psychiatric disorders makes investigating specific relationships between cognition and mental health, as well as their origins, difficult. It is likely that specific aspects of psychopathology will impact cognitive skills differently, and vice versa, specific cognitive profiles will lead to different psychopathological symptoms. A further challenge associated with investigating developmental effects is the heterogeneity in the average age of onset and diagnosis of different psychiatric conditions, which ranges widely, from childhood (e.g., ASD and ADHD) to adolescence (e.g., anorexia nervosa, anxiety, and depression) and early adulthood (e.g., schizophrenia and bipolar disorder).

One way of overcoming these challenges is to consider genetic risk. Genetic studies provide a powerful tool for uncovering and isolating shared and unique processes to each disorder. Research has shown that both cognitive abilities and psychiatric conditions are significantly heritable (8, 9), although heritability estimates vary widely across disorders, with some disorders, such as schizophrenia and ADHD, evincing higher heritability estimates than others, such as depression and anxiety (10–12). Studies have suggested that shared genetic factors might account for part of the associations between cognitive abilities and psychopathology. For instance, a twin study found that up to 40% of the observed association between symptoms of psychopathology and cognitive ability measured in early childhood could be attributed to shared genetic influences (13). Analyses of genome-wide association data also showed that genetic risk for psychopathology is significantly associated with cognition-related traits such as educational attainment (12).

Studies investigating the association between psychopathology and cognitive functioning have largely focused on the general psychopathology factor, also called the “p-factor” (14). The p-factor is thought to capture transdiagnostic effects across psychiatric conditions, and as such, it offers a useful framework for investigating generality and specificity in the development of the association between cognition and psychopathology. Recent findings suggest that the p-factor also operates at a genetic level (12, 15, 16), and that removing transdiagnostic genetic effects can reveal nuanced associations with other traits, including cognition (13, 17, 18). For instance, isolating transdiagnostic effects significantly changed the genetic correlations between psychiatric disorders and educational attainment, which increased significantly for schizophrenia and bipolar disorder and decreased significantly for depression (17). These findings highlight the importance of separating transdiagnostic from disorder-specific genetic factors to better understand the association between psychopathology and cognitive development.

Thus, in this study, we aim to investigate the complex relationship between psychopathology and cognitive development by leveraging genetic data to isolate transdiagnostic risk factors and explore generality and specificity of these associations. Using polygenic scores, which combine trait-associated genetic variants into a single composite index (19), in a large longitudinal sample, we investigate the association between transdiagnostic and disorder-specific genetic risk for psychopathology and cognitive development, from age 4 to 23. We examine how genetic risk for psychopathology is associated with differences in general cognitive ability, as well as with the development of verbal and non-verbal reasoning from childhood to early adulthood.

Finally, using family-level data, we explore the potentially different genetic pathways underlying the association between psychopathology and cognition. Investigating polygenic score associations considering difference between siblings allows us to look at relationships free from several potential confounding factors, including genetic (e.g., assortative mating, population stratification) and environmental influences shared by family members (e.g., passive gene-environment correlation) and therefore capture more direct genetic pathways.

Studies that have examined the association between genetic risk for psychopathology and the development of psychiatric symptoms have found that these effects largely operate through direct pathways. For example, studies have found a significant direct genetic association between genetic risk for externalizing behaviour and conduct problems/ADHD-related symptoms (20). as well as between genetic risk for neurodevelopmental disorders and related traits such as language, motor, social, and communication skills (21). However, these studies have primarily focused on within-trait prediction, while cross-trait associations—specifically, the pathways linking genetic risk for psychiatric disorders to cognitive profiles over development—remain unexplored. By integrating developmental and genetic frameworks, this study seeks to advance our understanding of the association between psychopathology and cognitive functioning.

## Methods and Materials

### Preregistration

The methods, hypotheses and analyses were preregistered on the Open Science Framework (https://osf.io/7enr5/).

### Sample

Participants were drawn from the Twins Early Development Study (TEDS) sample, a longitudinal study that recruited over 15,000 twin pairs born in England and Wales between 1994 and 1996. Currently, approximately 10,000 families remain actively involved in TEDS, nearly 30 years after the first wave of data collection. TEDS participants and their families are representative of the UK population in terms of ethnicity and socio-economic status for their birth cohort (22, 23).

This study used data collected across multiple waves of TEDS. Specifically, we analyzed data from 7 waves, when the twins were 4, 7, 9, 12, 16, 19 and 23 years old. Sample size varies between wares due to random drop-out and data availability (Supplementary Table 1). We excluded participants with severe medical conditions or uncertain/unknown zygosity from the analyses. TEDS received ethical approval from the research ethics committee of Kings College London (references: PNM/09/10–104 and HR/DP[20/21–22060). Written parental consent was obtained from all participants before data collection at every wave and from the twins at ages 16, 19 and 23.

### Measures

Here we provide a brief description of all the variables included in the present study. Please refer to https://www.teds.ac.uk/datadictionary for detailed descriptions of each sub-scale and items.

#### Measuring genetic risk for psychopathology using polygenic scores

In total, 23 psychopathology polygenic scores were created in N = 10,288 participants and used in the proposed analyses. The scores were calculated using three types of GWAS summary statistics: a) summary statistics from 11 GWAS of psychiatric disorders; b) summary statistics for a GWAS of a general psychopathology factor (‘p factor’; (17)); and c) 11 summary statistics of GWAS of psychiatric disorders, after statistically removing the p factor (‘non-p’ summary statistics; (17)).

The ‘non-p’ and p factor summary statistics were derived from a recent study investigating specificity in psychiatric disorders using Genomic Structural Equation Modelling (Genomic SEM) (17). The uncorrected polygenic scores were derived from the following 11 GWAS summary statistics: Attention deficit hyperactivity disorder (ADHD) (24), Autism spectrum disorder (ASD) (25), Post-traumatic stress disorder (PTSD) (26), Obsessive-compulsive disorder (OCD) (27), Anxiety (ANX) (28), Major Depressive Disorder (MDD) (29), Schizophrenia (SCZ) (30), Bipolar Disorder (BIP) (31), Anorexia Nervosa (AN) (32), alcohol dependence (ALCH) (33) and Tourette syndrome (TS) (34).

Polygenic scores were created using LDPred2 in the R package bigsnpr (35). Compared to other methods and previous iterations (i.e., LDPred1), LDPred2 offers higher predictive power and overcomes previous limitations (35). A standardized pipeline was applied to create all polygenic scores. Genotyping information can be found in the Supplementary Methods.

Standard genetic confounding variables, including the top 10 principal components of genetic ancestry, genotyping batch, sex, and age, were included in all analyses.

#### Measures of Cognitive Abilities

Cognitive abilities were measured using standardized cognitive tests collected at ages 4, 7, 9, 12, 16, 19 and 23 years. These composite scores were designed to accommodate children’s developmental stages and were derived from different questionnaires at each age. At each time point, three composite scores were calculated: verbal cognitive ability, nonverbal cognitive ability, and general cognitive ability (g). Composite scores for verbal and nonverbal reasoning were derived from the mean of the standardized tests described below. The standardized g composite was derived by taking the mean of the standardized verbal and nonverbal composite scores.

**Age 4.** Cognitive abilities were assessed using four tests administered as booklets sent to parents to complete. Two tests assessed verbal reasoning: 48 item MacArthur Communicative Development Inventories (MCDI) (36) and an ordinal grammar composite scale. Nonverbal reasoning was assessed using the parent-report and parent-administered Parental Report of Children’s Ability questionnaire (PARCA), which was developed by the TEDS team based on a battery of scales (PARCA; (37))

**Age 7.** Cognitive ability was measured using four child-completed tests administered over the telephone by trained research assistants, including two verbal tests (a 13-item similarity test and 18-item vocabulary test) both derived from the Wechsler Intelligence Scale for Children (WISC-3) (38) and two nonverbal tests (a 9-item conceptual groupings test from McCarthy scales (39) and a 21-item WISC picture completion test (38).

**Age 9.** Cognitive ability was assessed using four tests that were administered as booklets sent to families to be completed by the children. Verbal reasoning was assessed using two tests, the first 20 items of words test and the first 18 items of a general knowledge test from WISC-3-PI (40). Nonverbal reasoning was assessed using the shapes and puzzle tests from the cognitive abilities test 3rd edition (CAT3) (41) figure classification and figure analogies.

**Age 12**. Cognitive ability was assessed using four tests that were administered online and completed by the children. Verbal reasoning was assessed using the 30-item full version of the word test and the general knowledge test from WISC-3-PI (40). Nonverbal reasoning was assessed using the pattern test adopted from the Raven’s standard progressive matrices (42) and the picture completion test adopted from the WISC-3-UK (43).

**Age 16.** Cognitive ability was assessed using two tests administered online and completed by the children. Verbal reasoning was assessed using the Mill Hill vocabulary test (43) and nonverbal reasoning was assessed using the Raven’s standard progressive matrices test (42).

**Age 19.** Spatial abilities are measured separately. They were administered when the sample aged 19-22 through two online batteries: ‘King’s Challenge’ and ‘Spatial Spy’. The first battery measures spatial manipulation, while the second focused on spatial navigation. Details of the battery development and its tests were outlined elsewhere (44, 45).

**Age 23.** Cognitive skills were measured using Pathfinder (46), a brief, gamified measure of general, verbal and nonverbal cognitive abilities developed and validated by the TEDS team.

#### Additional Measures

Four cognition-related PGSs and the family socio-economic status (SES) were included in robustness checks analyses (See Statistical Analysis section). The four cognitive PGSs were: cognitive and non-cognitive abilities (47), a genomic general cognitive ability factor (genomic g; (48)), and intelligence (49). The four cognitive polygenic scores were selected based on their power and representativeness.

Family SES was measured using a composite of five variables: mother and father employment levels, mother and father educational levels, and mother’s age on birth of first child. The SES composite score was the most used SES variable in the TEDS and showed great stability across development (22).

### Statistical Analysis

#### Regression of Cognitive Abilities on Polygenic Scores

Multiple regression was used to test for predictions of psychopathology polygenic scores on cognitive ability scores. We used the full sample in our analysis and accounted for non-independence of observations for DZ co-twins using the generalized estimating equation (GEE). FDR correction (50) was applied to adjust the criteria for significance. Differences in PGS predictions before and after correcting for p, and between verbal and nonverbal cognitive abilities, were tested for significance using z score test.

#### Within-family and Between-family Analyses

To further interpret the associations and explore the pathways linking genetic risk for psychopathology and cognitive outcomes, we applied a twin difference design. Twin difference analyses rely on how, through meiosis, sibling differences in genotypes are randomized, meaning that siblings have an equal probability of inheriting any given parental allele. As a result, within-sibling polygenic score associations are not influenced by genetic and environmental factors that operate between families such as passive gene-environment correlation, assortative mating and population stratification (51, 52).

#### Robustness Check

We re-ran the multiple regression analyses with additional covariates to test the robustness of the results, including family socio-economic status (SES) and cognitive polygenic scores (see Measures section). Each robustness check variable was included in the multiple regression analysis separately.

#### Cognitive Change Score Analysis

In the supplementary note, we present the method and results of our pre-registered analysis examining cognitive change scores of general, verbal and nonverbal abilities.

## Results

### Psychopathology and General Cognitive Abilities

Figure 1 presents the results of the multiple regression analyses examining the association between genetic risk for psychopathology, measured using polygenic scores, and general cognitive ability from age 4 to age 23 years. Transdiagnostic genetic risk, reflected in the p factor polygenic score, was minimally associated with general cognitive ability across development. The only significant association was observed at age 23 (β = –0.060, SE = 0.020, p_fdr_ = 0.012). Consequently, the effect sizes of the predictions remained virtually unchanged when comparing disorder-specific polygenic scores before and after accounting for transdiagnostic effects (see Figure 1 blue vs. yellow bars).

**Figure 1.**
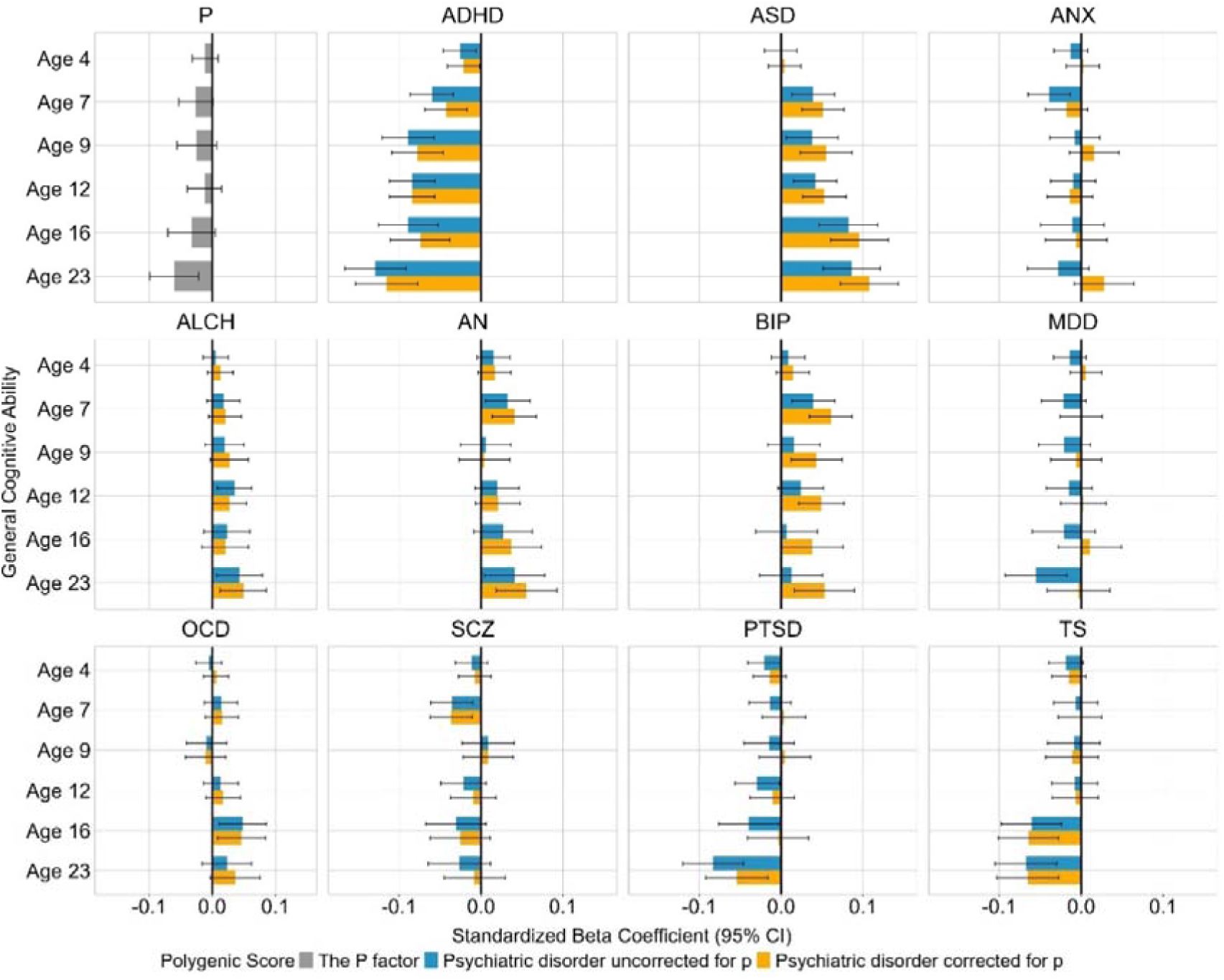
Associations between polygenic scores for psychiatric disorders and general cognitive ability across development. The grey bars represent the associations (standardized beta coefficient) between transdiagnostic genetic risk for psychopathology (p) and general cognitive ability over development, from age 4 to 23. The blue and yellow bars show associations between genetic risk for each psychiatric disorder and general cognitive ability over development before (blue) and after (yellow) removing the effects of transdiagnostic genetic risk. The error bars indicate 95% confidence intervals. ADHD = Attention deficit hyperactivity disorder, ASD = Autism spectrum disorder, PTSD = Post-traumatic stress disorder, OCD = Obsessive-compulsive disorder, ANX = Anxiety, MDD = Major Depressive Disorder, SCZ = Schizophrenia, BIP = Bipolar Disorder, AN = Anorexia Nervosa, ALCH = Alcohol dependence and TS = Tourette syndrome.

Associations between individual psychiatric disorders and general cognitive ability were developmentally consistent for each disorder and, for some disorders (e.g., ADHD and ASD) effect sizes increased across development. Particularly, the ADHD polygenic score showed the strongest association across development, ranging from β= –0.026, SE = 0.010, p_fdr_ = 0.038 at age 4 to β = –0.129, SE = 0.0192, p_fdr_ < 0.001 at age 23. The ASD polygenic score also showed significant associations with general cognitive ability across development (ranging between β = 0.039, SE = 0.013, p_fdr_ = 0.015 at age 7 and β = 0.086, SE = 0.018, p_fdr_ < 0.001 at age 23), except for early childhood (β =-0.0007, SE = 0.010, p_fdr_ = 0.95).

Interestingly, associations between genetic risk for psychopathology and general cognitive ability were negative for some disorders (e.g., ADHD, Tourette’s and PTSD) and positive for others (e.g., ASD, anorexia nervosa and bipolar), however, some did not survive FDR correction (see Supplementary Table 2). Robustness checks showed that, most associations between psychiatric polygenic scores and general cognitive ability remained significant after controlling for family socio-economic status and cognition-related polygenic scores by means of multiple regression (Supplementary Table 2).

### Psychopathology and Specific Cognitive Abilities

To further explore the association between psychopathology and cognition, we extended our analyses to examine the prediction from psychiatric polygenic scores to three more specific subcomponents of cognitive functioning: verbal reasoning, nonverbal reasoning and spatial ability. Figure 2 presents the psychiatric polygenic scores prediction of verbal and nonverbal reasoning (see Supplementary Figure 3 for the same results using psychiatric polygenic scores corrected for p). Overall, most disorders, except for anxiety, alcohol dependence and obsessive-compulsive disorder, showed at least one significant association with either verbal or nonverbal reasoning. Similar to the what observed for general cognitive ability, removing transdiagnostic genetic risk did not significantly alter psychiatric disorder-cognition associations (See Supplementary Table 7).

**Figure 2.**
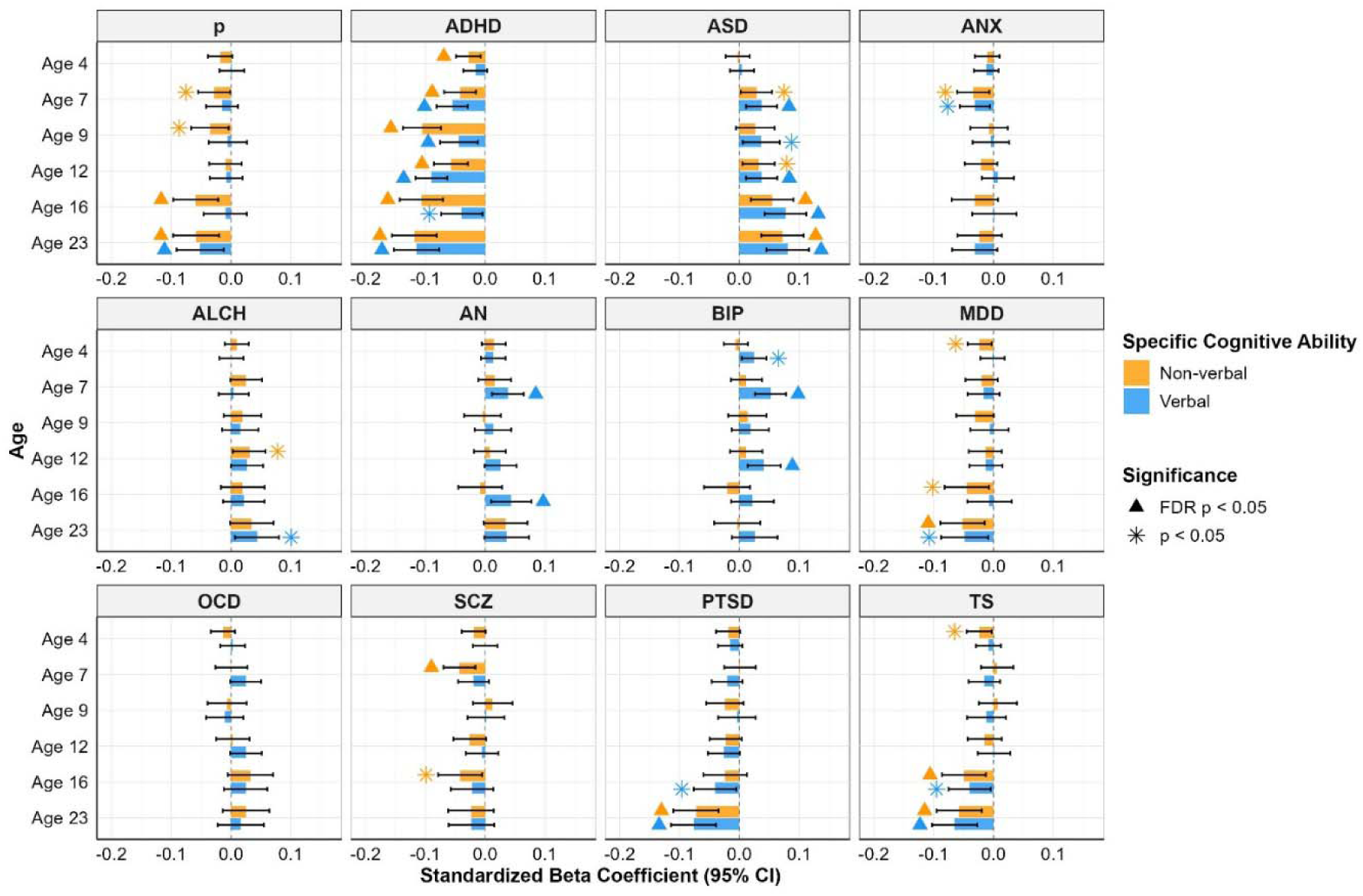
Associations between polygenic scores for psychiatric disorders and verbal and nonverbal reasoning across development. The x-axis shows the standardized beta coefficient, and the y-axis cognitive outcome at ages 4 to 23 years. Each panel shows results for a different psychiatric disorder. The yellow bars are associations with nonverbal reasoning and the blue bars with verbal reasoning. Error bars indicate 95% confidence intervals. The stars indicate significance at threshold p < 0.05 and the triangles significance after FDR correction. ADHD = Attention deficit hyperactivity disorder, ASD = Autism spectrum disorder, PTSD = Post-traumatic stress disorder, OCD = Obsessive-compulsive disorder, ANX = Anxiety, MDD = Major Depressive Disorder, SCZ = Schizophrenia, BIP = Bipolar Disorder, AN = Anorexia Nervosa, ALCH = Alcohol dependence and TS = Tourette syndrome.

**Figure 3.**
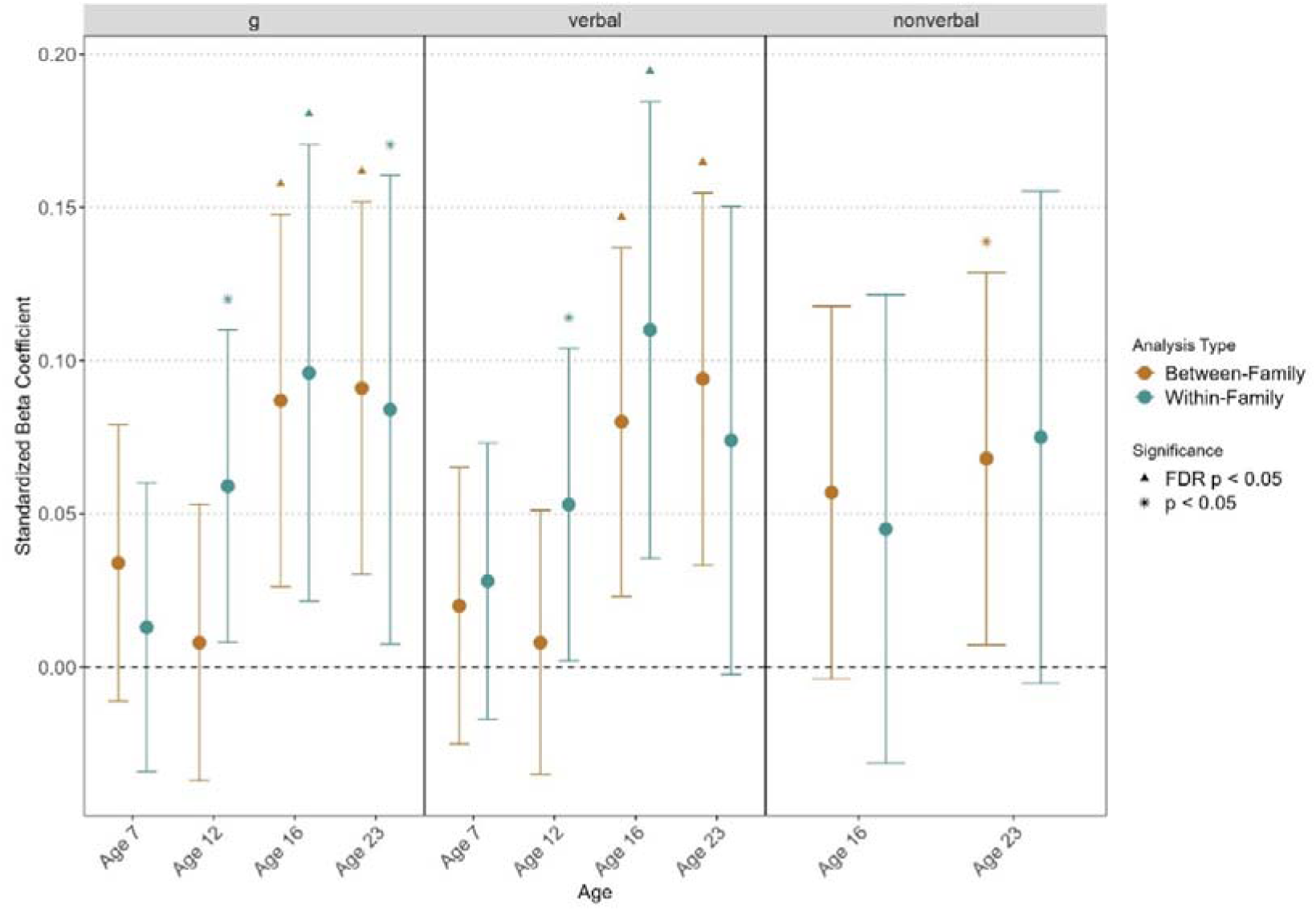
Family-level analyses of the association between autism spectrum disorder and general cognitive ability, verbal and nonverbal reasoning. This plot presents the within-family analyses for the significant associations identified for the ASD polygenic score. The brown dots represent between-family effects of the ASD polygenic scores on cognitive outcomes, and the green dots indicate within-family effects. The error bars indicate 95% confidence intervals. The stars represent significance at a p < 0.05 threshold, and the triangles significance after FDR correction.

However, when considering greater specificity at the level of cognitive skills, a clearer differentiation between the p-factor and disorder-specific effects began to emerge. The p-factor polygenic score exhibited a stronger association with nonverbal reasoning across multiple ages (β = –0.059, SE = 0.019, p_fdr_ = 0.018 at age 16 and β = –0.058, SE = 0.020, p_fdr_ = 0.021 at 23), while its association with verbal reasoning was only significant in early adulthood (β = –0.052, SE = 0.02, p_fdr_ = 0.046). In contrast, polygenic scores associated with individual disorders—including those for ADHD, ASD, anorexia nervosa, and bipolar disorder—displayed either stronger associations with verbal reasoning (e.g., anorexia nervosa and bipolar disorder) or significant links across all three cognitive domains across development (e.g., ADHD, autism, PTSD).

When comparing the strength of the polygenic score predictions for verbal and nonverbal reasoning, we found that ADHD was more strongly associated with nonverbal reasoning at ages 9 and 16, while bipolar disorder and anorexia nervosa were more significantly associated with verbal abilities (see Supplementary Figure 2), although some of those differences did not pass FDR correction (Supplementary Table 9). For other disorders—including ASD, anxiety, depression, and schizophrenia—trends favouring either verbal or nonverbal abilities were observed but the difference was not significant (Figure 2).

Regarding spatial abilities, spatial manipulation ability was significantly predicted by the polygenic scores for schizophrenia, anorexia nervosa, ASD, and ADHD (Supplementary Figure 4), though only the association between ADHD, ASD and schizophrenia passed FDR correction (Supplementary Table 5). Spatial navigation was significantly associated with genetic risk for ASD, PTSD and schizophrenia but only the schizophrenia association remained significant after FDR correction (Supplementary Table 5).

## Discussion

This study examined the associations between genetic risks for psychopathology and cognitive development, from early childhood to early adulthood. By leveraging polygenic scores derived from genome-wide association studies, we tested whether genetic risk for different psychiatric disorders, before and after we removed transdiagnostic effects, was associated with specific profiles of cognitive functioning over development. First, our results indicate that, contrary to what has been observed for the development of psychiatric symptoms, which are best predicted by transdiagnostic effects (53), the association between psychopathology and general cognitive ability is driven by disorder-specific genetic risk rather than transdiagnostic influences. Second, we found that genetic risk for different psychiatric disorders is differentially associated with specific cognitive profiles, with some disorders (e.g., bipolar disorder and anorexia nervosa) more closely linked to verbal reasoning while others (e.g., ADHD) to non-verbal cognitive skills. Third, within family analyses highlighted how these population-level associations capture a mixture of direct and indirect effects, with ASD-related genetic risk showing the most robust direct effects. These findings provide us with a deeper understanding of how genetic risk for psychiatric disorders is linked to the development of specific cognitive profiles and highlight how research into cognitive functioning in psychopathology would gain from focusing on individual disorders rather than transdiagnostic factors.

The first main finding of the study was that genetic risk linked to the transdiagnostic p-factor had limited predictive value for general cognitive abilities, with significant associations emerging only in early adulthood. In contrast, genetic vulnerability to individual psychiatric disorders predicted general cognitive ability, both before and after controlling for the p-factor. These associations were consistently observed across development, and effect sizes increased over time. However, disorder–cognition associations were not uniform across disorders. For some conditions, such as ADHD, PTSD, and Tourette’s syndrome associations were consistently negative whereas for other disorders such as ASD, alcohol dependence, anorexia nervosa, and bipolar disorder they were consistently positive. This suggests that genetic risk related to some psychiatric conditions might confer potential cognitive advantages while genetic risk associated with others potential disadvantages.

These results are in line with previous findings on the association between psychiatric disorders and cognition. For example, ADHD has been extensively studied, with literature documenting that children and adults with ADHD achieve lower scores in cognitive tests across clinical (54, 55) and subclinical (56, 57) samples. A genetically informative study using a general population sample also found that polygenic risk for ADHD was negatively associated with performance IQ, even after controlling for a latent neurodevelopmental risk score (58). Deficits in cognitive functioning have also been associated with PTSD, beyond the effects of other psychiatric symptoms such as depression (59) and beyond the influence of traumatic experiences (60). In the case of Tourette’s syndrome, although findings are less consistent (61), negative associations with general cognitive ability have been reported (62), especially in population-based samples (63).

On the other hand, for ASD, our results highlight its positive genetic correlation with general cognitive ability (17, 64) and are in line with a previous meta-analysis of the association between the ASD polygenic score and general cognitive ability across three independent cohorts (65). Though the literature often reports negative (66) or no associations between bipolar disorder and cognitive ability (67, 68), it has been reported that genetic liability toward bipolar disorder is associated with skills related to cognitive outcomes such as creativity (69, 70), educational attainment (71), and a lower risk of cognitive deficits (72). Genetic liability to anorexia nervosa was positively associated with cognitive performance (17) and our findings are in line with research that does not consider genetic risk (73).

These results suggest that cognitive characteristics related to psychopathology differ for different psychiatric disorders and that disorder-specific genetic factors play a more substantial role in shaping cognitive outcomes than transdiagnostic effects. As such, rather than focussing on investigating relationships between transdiagnostic genetic risk and cognition (13, 74–76), studies would benefit from investigating these relationships separately for each psychiatric disorder. This contrasts with findings on genetic vulnerability to psychiatric symptoms, where transdiagnostic genetic effects provide the strongest predictive power across both diagnostic categories and symptom continua (17, 53, 77). Our findings highlight the complexity of psychiatric disorders and their associated differences in cognitive functioning.

Our results also revealed nuanced associations between psychiatry disorders and cognitive domains. For instance, many disorders demonstrated robust predictive effects on specific cognitive abilities, with the ADHD polygenic score showing the strongest negative association with non-verbal reasoning. In contrast, the anorexia nervosa and bipolar disorder polygenic scores were more positively associated with verbal abilities across development. In term of spatial cognitive ability, only ADHD and schizophrenia were negatively associated with spatial manipulation abilities. These results reflect previous clinical findings that individuals with ADHD perform significantly lower in spatial and non-verbal abilities if compared to healthy controls, including domains such as visuo-spatial perception and abstract thinking (78), working memory (79), and processing speed (80), where inattention may contribute to poorer performance on tasks with a higher cognitive load. A meta-analysis of 26 studies also showed that ADHD patients had greater deficits in visual-spatial storage and manipulation skills when compared with verbal reasoning (81). On the other hand, genetic vulnerability to autism was found to be equally linked to verbal and nonverbal reasoning. In line with our findings, previous studies have found that verbal and nonverbal reasoning equally contributed to language outcomes in children with ASD (82), and that adults with ASD demonstrated homogeneous cognitive profiles for verbal and nonverbal reasoning, irrespective of the severity of their ASD symptoms (83).

At the family level, we found that the pathways through which genetic risk for psychiatric disorders contributes to cognitive development vary across disorders. For ADHD, alcohol dependence, anorexia nervosa, PTSD, and Tourette’s syndrome, the effects were primarily operating through between-family pathways. In contrast, ASD, bipolar disorder, and schizophrenia exhibited both within– and between-family effects. This is in line with previous research that found evidence for direct genetic effects on neurodevelopmental symptoms in early childhood (20, 21). This distinction underscores the heterogeneity in how different disorders contribute to cognitive outcomes and highlights the importance of disentangling different genetic and environmental pathways developmentally (52, 84, 85). It also suggests that disorders may be differentially influenced by individual– and population-level factors, therefore interventions aimed at mitigating the impact of psychopathology on cognitive functioning, and vice versa, should consider these different exposures as well as specific vulnerability.

Several limitations should be acknowledged. First, although TEDS is representative of the population of England and Wales for their birth cohort, it is not representative of the current population. For example, the TEDS families score higher on socio-economic outcomes compared to the current population mean. This is particularly important as socio-economic outcomes have been linked to cognitive development (86) and mental health (87), and it limits the generalizability of our findings to broader populations and new generations of children. Second, while our study allowed us to view the association between psychopathology and cognition from a genetic and developmental perspective, we did not model relationships longitudinally due to different measures available at different ages.

Future studies using longitudinal methods such as latent growth models or random intercept cross-lag models might investigate the dynamic relations between cognitive functioning and symptoms of psychopathology, although this presents other challenges, for example differences in the age of onset of psychiatric conditions (88). Third, our cognitive measures, while comprehensive, might not fully capture the multidimensional nature of cognitive development (89). Incorporating further specific measures such as tests of executive functioning and memory could provide a more nuanced picture (90). Fourth, it should also be acknowledged that polygenic score effects were small, particularly when decomposing association into between– and within-family associations. Fifth, we acknowledge that, while we modelled transdiagnostic effects using a common factor model, the p factor has been modelled differently, for example using a hierarchical factor model (12, 18). However, this is unlikely to have substantially affected the main findings, as transdiagnostic effects were found to play only a marginal role in their association with cognitive development.

## Data Availability

All data produced in the present study are available upon reasonable request to the authors

## Acknowledgements

We gratefully acknowledge the ongoing contribution of the participants in the Twins Early Development Study (TEDS) and their families. TEDS is supported by the UK Medical Research Council (MR/V012878/1 and previously MR/M021475/1. MM is supported by a Jacobs Foundation Research Fellowship (#2024-1533-00) and UK Medical Research Council Grant UKRI1506. WL is supported by a Chinese Scholarship Council PhD scholarship. EK is supported by a PhD scholarship awarded by the Economic and Social Research Council.

## Author information

WL and MM conceived and designed the study. WL analysed the data. MM supervised the work. WL and MM wrote the paper with helpful contributions from EK, AA, KR, and RP. All authors contributed to the interpretation of the data, provided critical feedback on paper drafts and approved the final draft.

## Supplementary Note: Pre-registrated cognitive change score analysis

This supplementary note outlines the pre-registered analysis conducted to examine cognitive change scores and their association with genetic risk for psychopathology. The objective was to investigate how genetic predispositions to psychiatric conditions relate to cognitive development from childhood to adulthood.

Using the composite cognitive measures described in the main text, we calculated cognitive change scores for general cognitive ability, verbal, and nonverbal reasoning. These scores represented an individual’s relative cognitive development from childhood to adulthood within the cohort. Specifically, cognitive change scores were computed by subtracting standardized cognitive scores at age 7 from standardized scores at age 21. Participants with change scores more than one standard deviation (SD) below the cohort mean were classified as having slower cognitive development, whereas those with scores more than one SD above the mean were classified as having faster development relative to their peers. Sample sizes for each group across the three domains are presented in Supplementary Table 1.

Participants were divided into two groups—higher cognitive change and lower cognitive change—based on their scores being *≥*1 SD above or below the cohort mean. Regression analyses between the genetic risks of psychopathology and cognitive change scores were conducted separately for each group to test whether the relationship between polygenic risk scores for various psychiatric disorders and cognitive change scores varied by developmental trajectory.

The regression results are shown in Supplementary Table 6. In summary, the analyses revealed limited significant associations, which could be due to small sample size in each group. For general cognitive ability, polygenic scores for obsessive-compulsive disorder and Tourette’s syndrome were significantly associated with cognitive change only in the faster development group. In contrast, verbal reasoning change scores were predicted by depression and Tourette’s polygenic scores in the slower development group, and by PTSD polygenic scores in the faster development group. No polygenic scores significantly predicted nonverbal reasoning changes in either group. However, none of these associations survived false discovery rate (FDR) correction.

**Figure S1:**
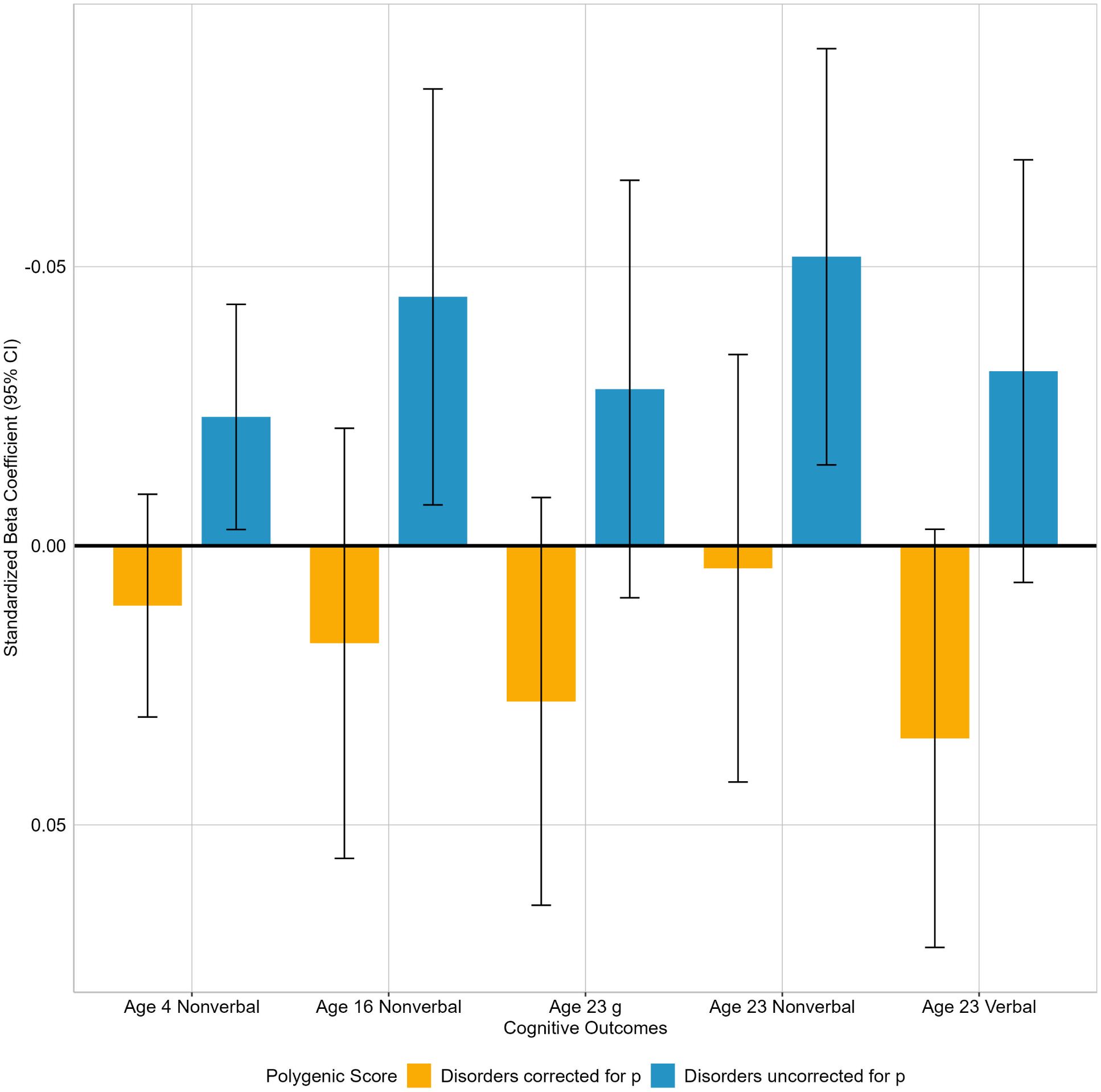
Significant differences in cognitve ability prediction of psychiatry disorders before and after controlling for the transdiagnostic p factor.

The figure shows the significant differences in cognitve ability prediction of psychiatry disorders before and after controlling for the transdiagnostic p factor. The differences are statistically significant at the 0.05 level.

**Figure S2:**
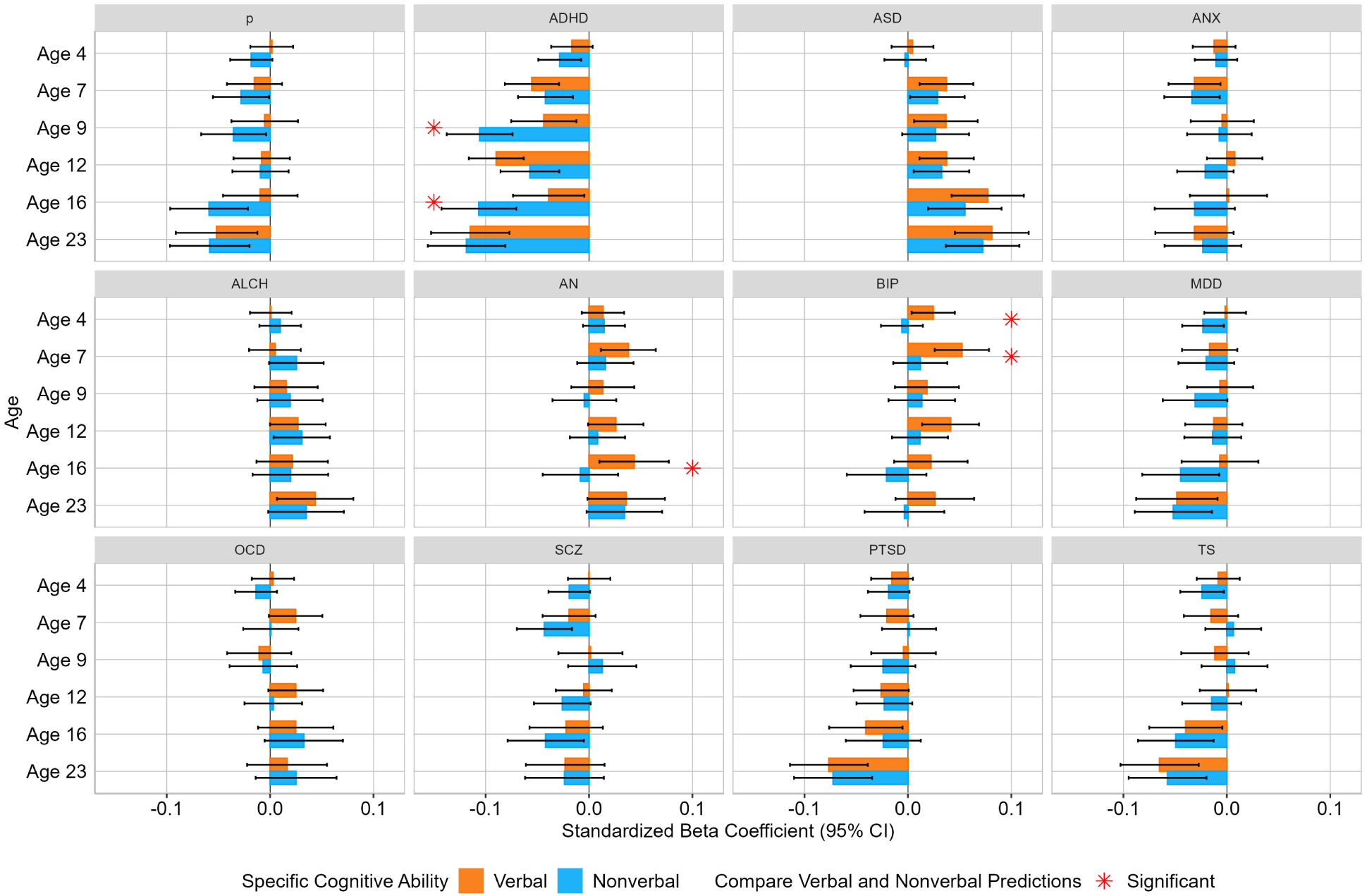
Significant Differences in Associations Between Verbal and Non-Verbal Cognitive Ability and Psychiatric Polygenic Scores.

The figure shows the comparison between verbal and non-verbal cognitive ability for each psychaitric disorder PGS uncorrected for p factor. The differences are statistically significant at the 0.05 level.

**Figure S3:**
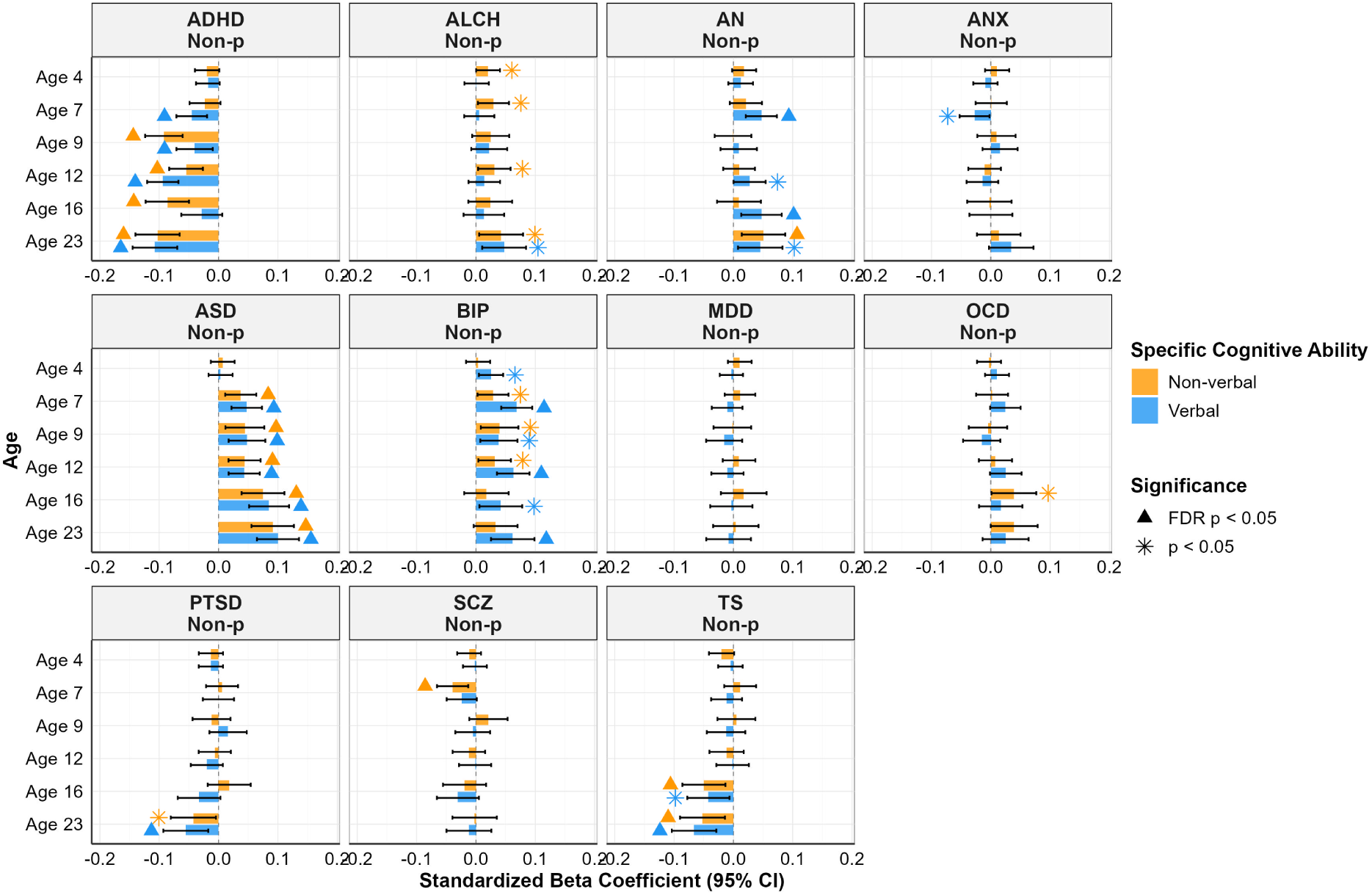
Associations Between Verbal and Non-Verbal Cognitive Ability and Psychiatric Polygenic Scores after Correcting for p factor.

The figure shows the associations between specific cognitive ability and psychiatric polygenic scores after correcting for p factor.

**Figure S4:**
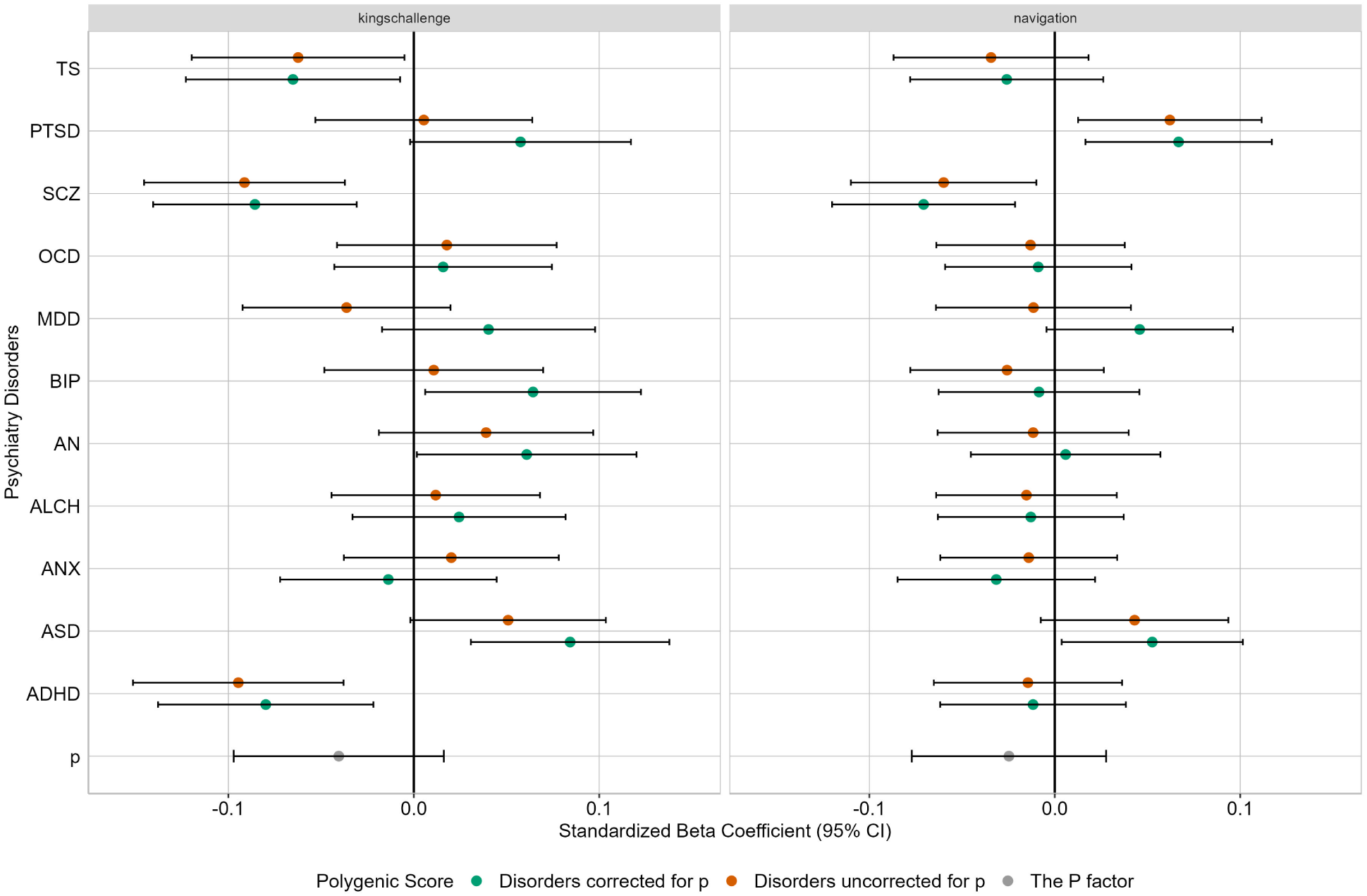
Spatial Cognitive Ability with Psychiatric Polygenic Scores before and after correcting for p factor.

The figure illustrates spatial cognitive abilities and their associations with psychiatric polygenic scores, comparing results before and after correcting for the transdiagnostic p factor.

**Figure S5:**
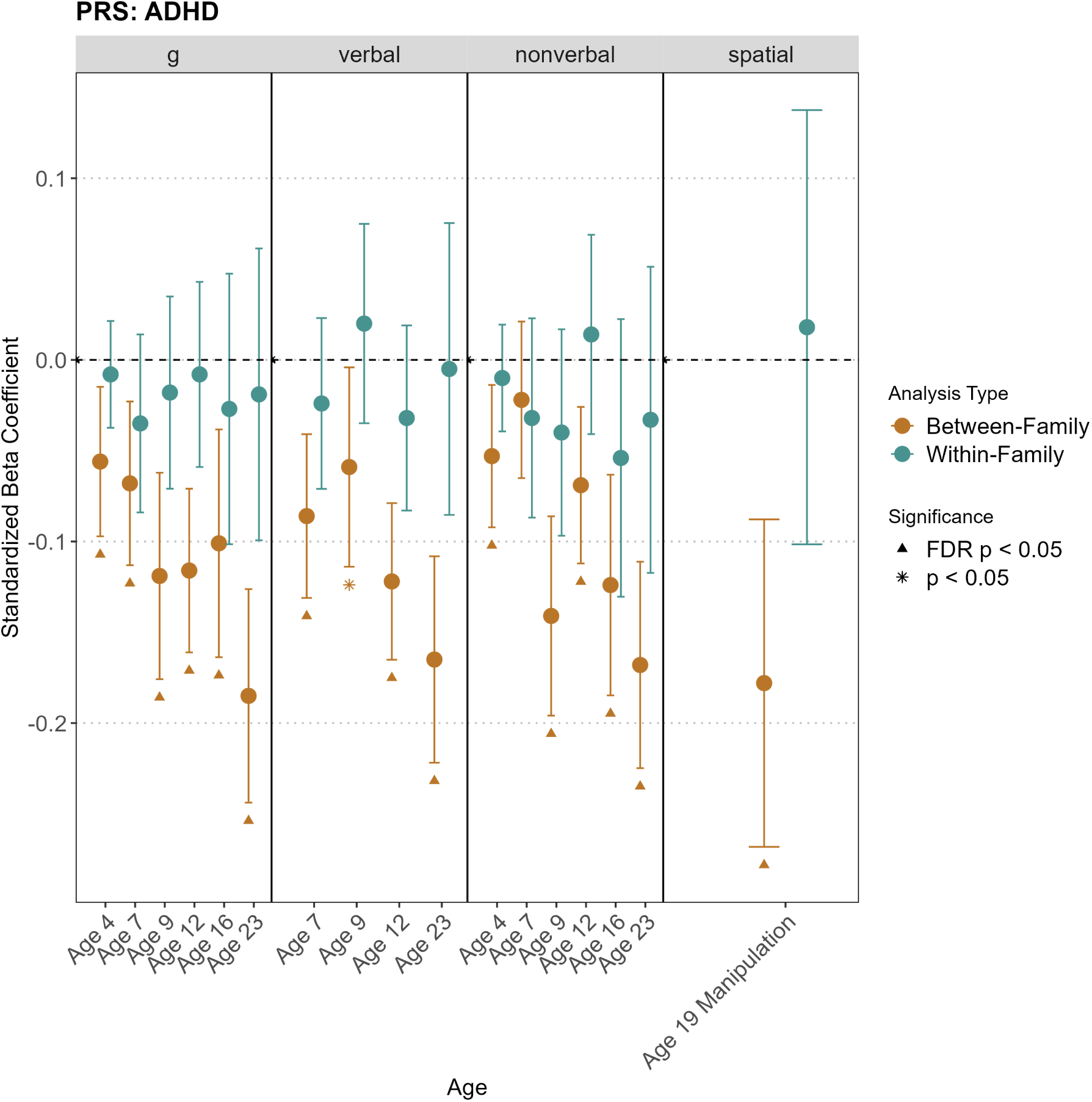
Direct and Indirect Genetic Effects of genetic risk of ADHD on Cognitive Abilities.

The figure presents the direct genetic effects and family-mediated indirect genetic effects of ADHD polygenic scores on cognitive abilities across development.

**Figure S6:**
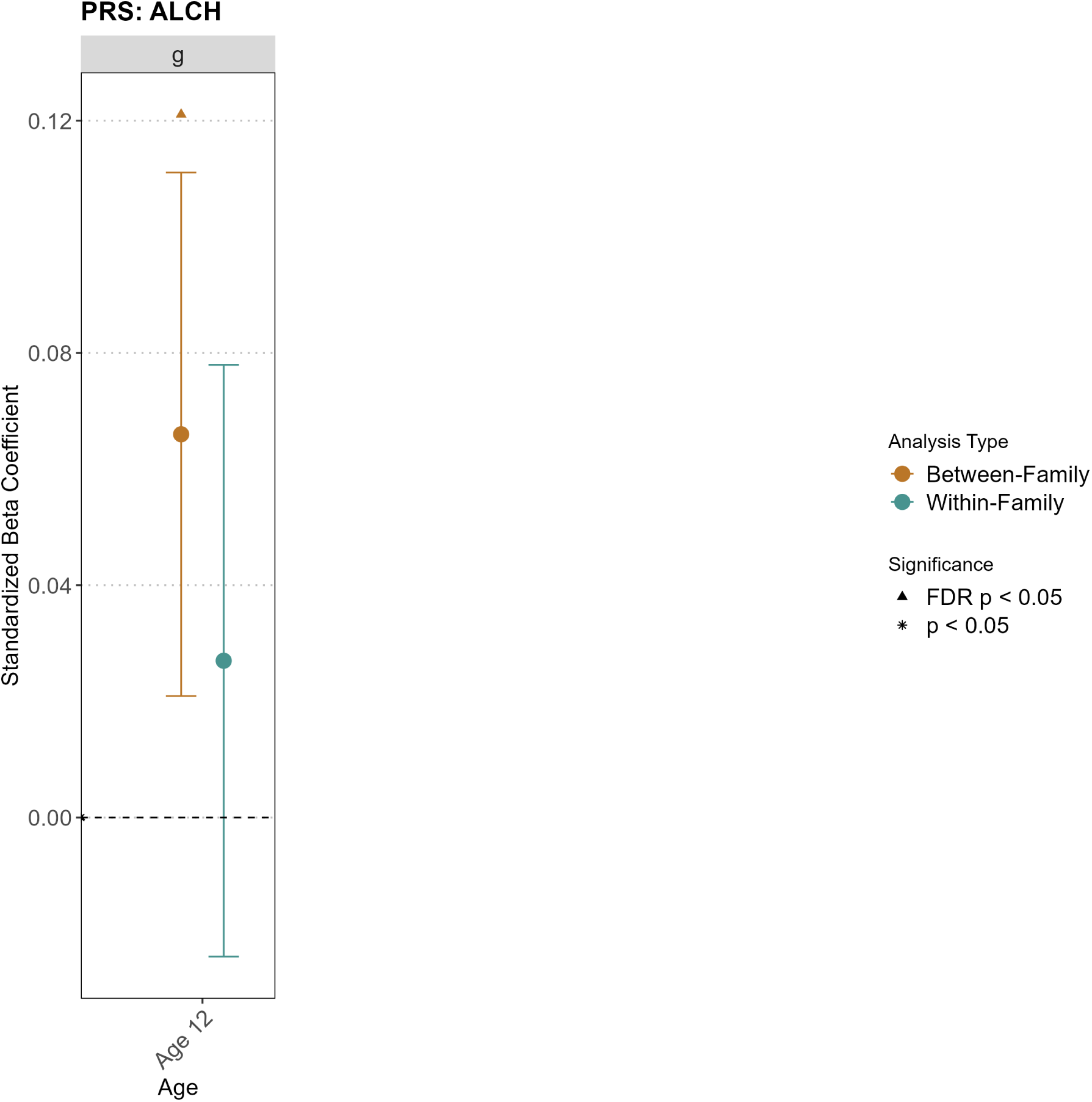
Direct and Indirect Genetic Effects of genetic risk of ALCH on Cognitive Abilities.

The figure presents the direct genetic effects and family-mediated indirect genetic effects of alcohol use disorder polygenic scores on cognitive abilities across development.

**Figure S7:**
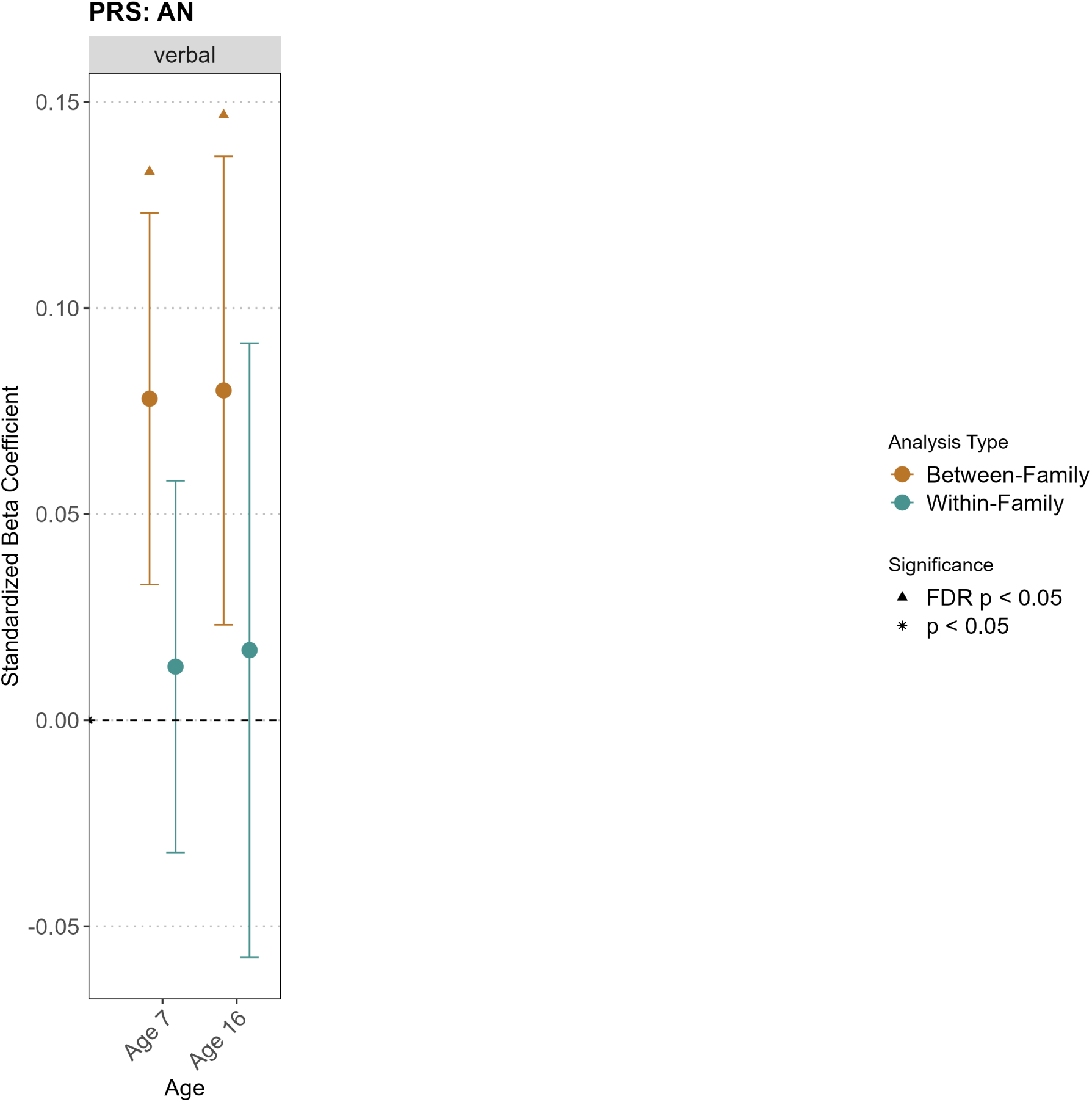
Direct and Indirect Genetic Effects of genetic risk of AN on Cognitive Abilities.

The figure presents the direct genetic effects and family-mediated indirect genetic effects of anorexia nervosa polygenic scores on cognitive abilities across development.

**Figure S8:**
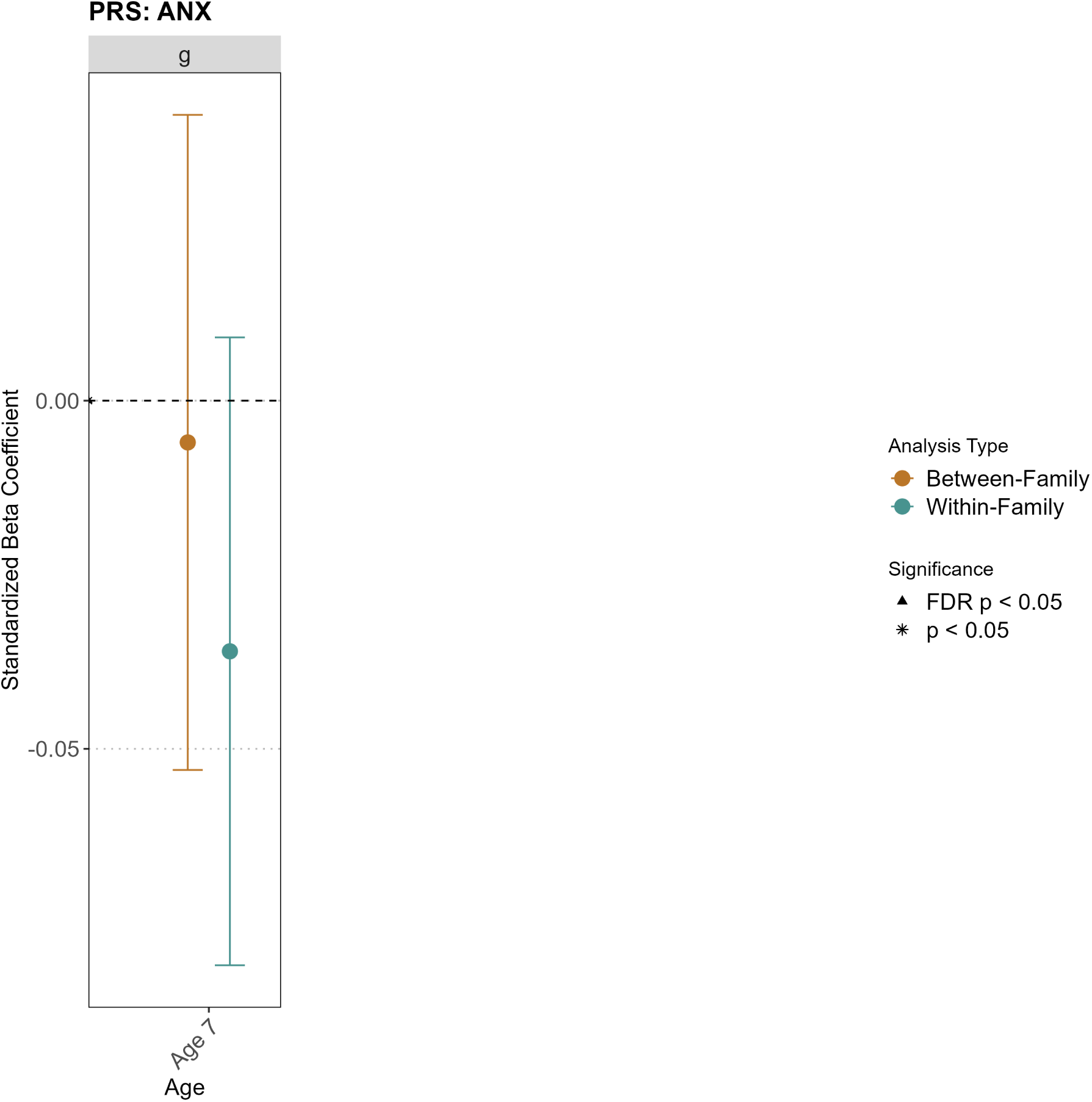
Direct and Indirect Genetic Effects of genetic risk of ANX on Cognitive Abilities.

The figure presents the direct genetic effects and family-mediated indirect genetic effects of anxiety disorder polygenic scores on cognitive abilities across development.

**Figure S9:**
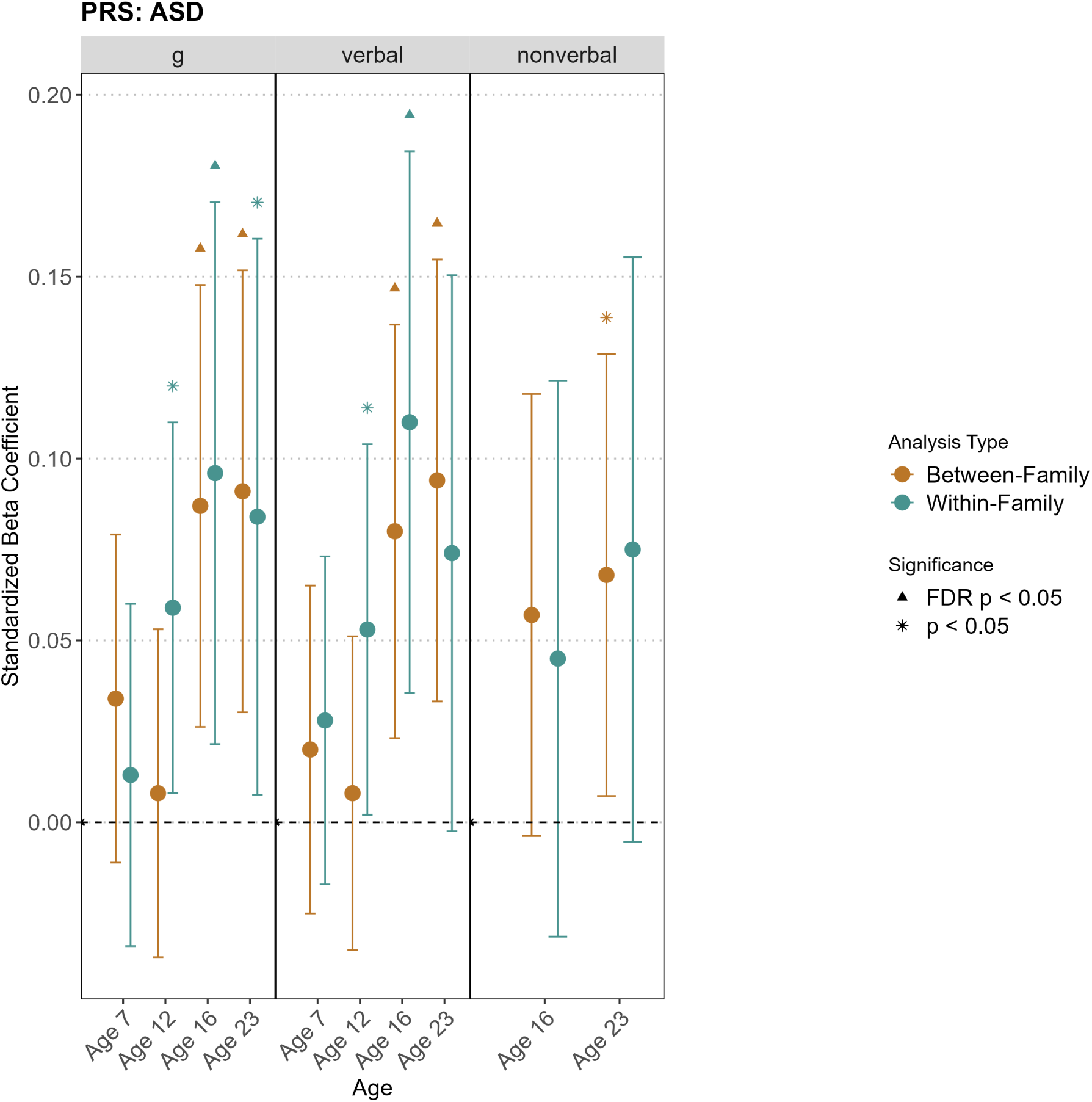
Direct and Indirect Genetic Effects of genetic risk of ASD on Cognitive Abilities.

The figure presents the direct genetic effects and family-mediated indirect genetic effects of autism spectrum disorder polygenic scores on cognitive abilities across development.

**Figure S10:**
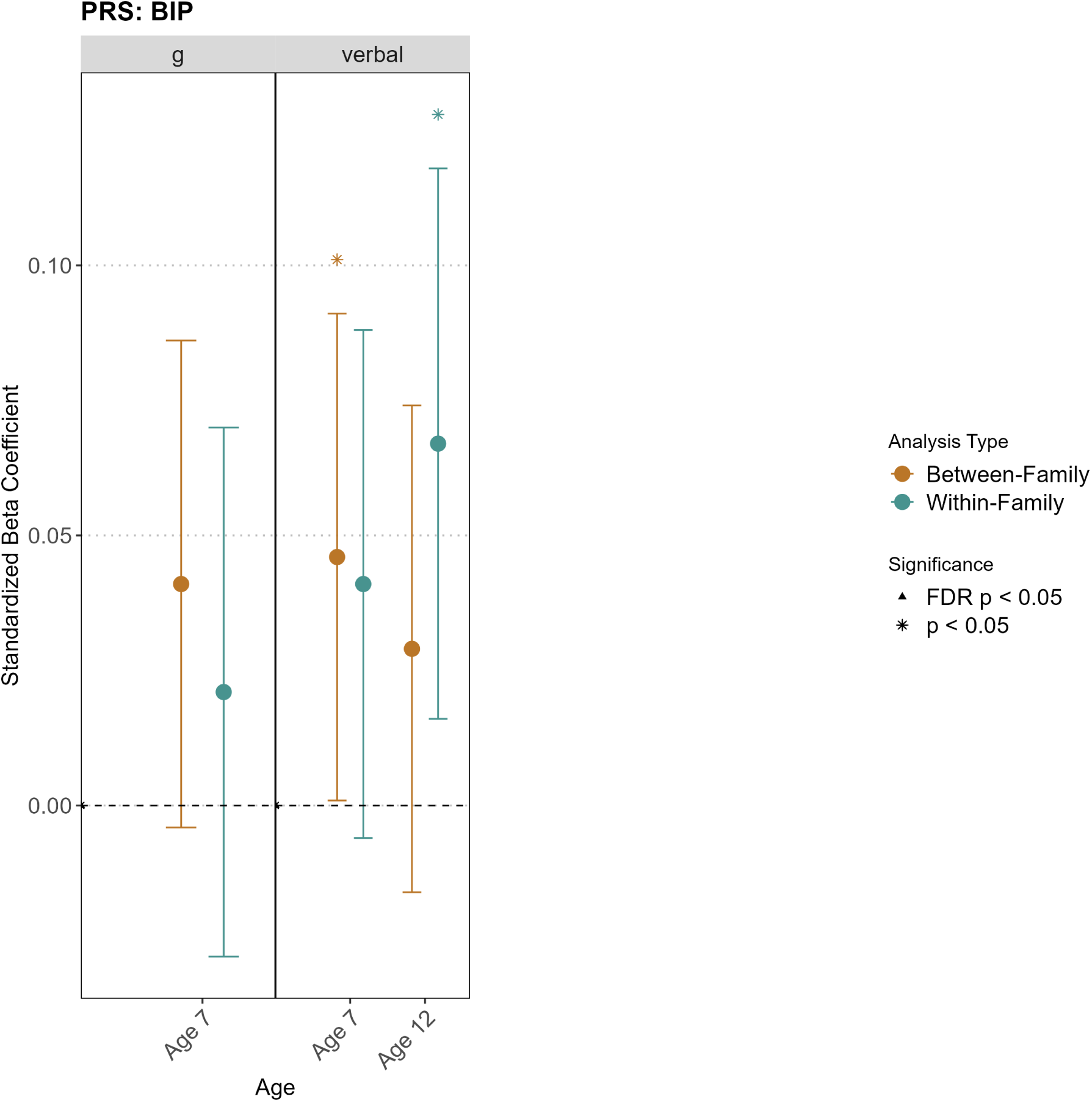
Direct and Indirect Genetic Effects of genetic risk of BIP on Cognitive Abilities.

The figure presents the direct genetic effects and family-mediated indirect genetic effects of bipolar disorder polygenic scores on cognitive abilities across development.

**Figure S11:**
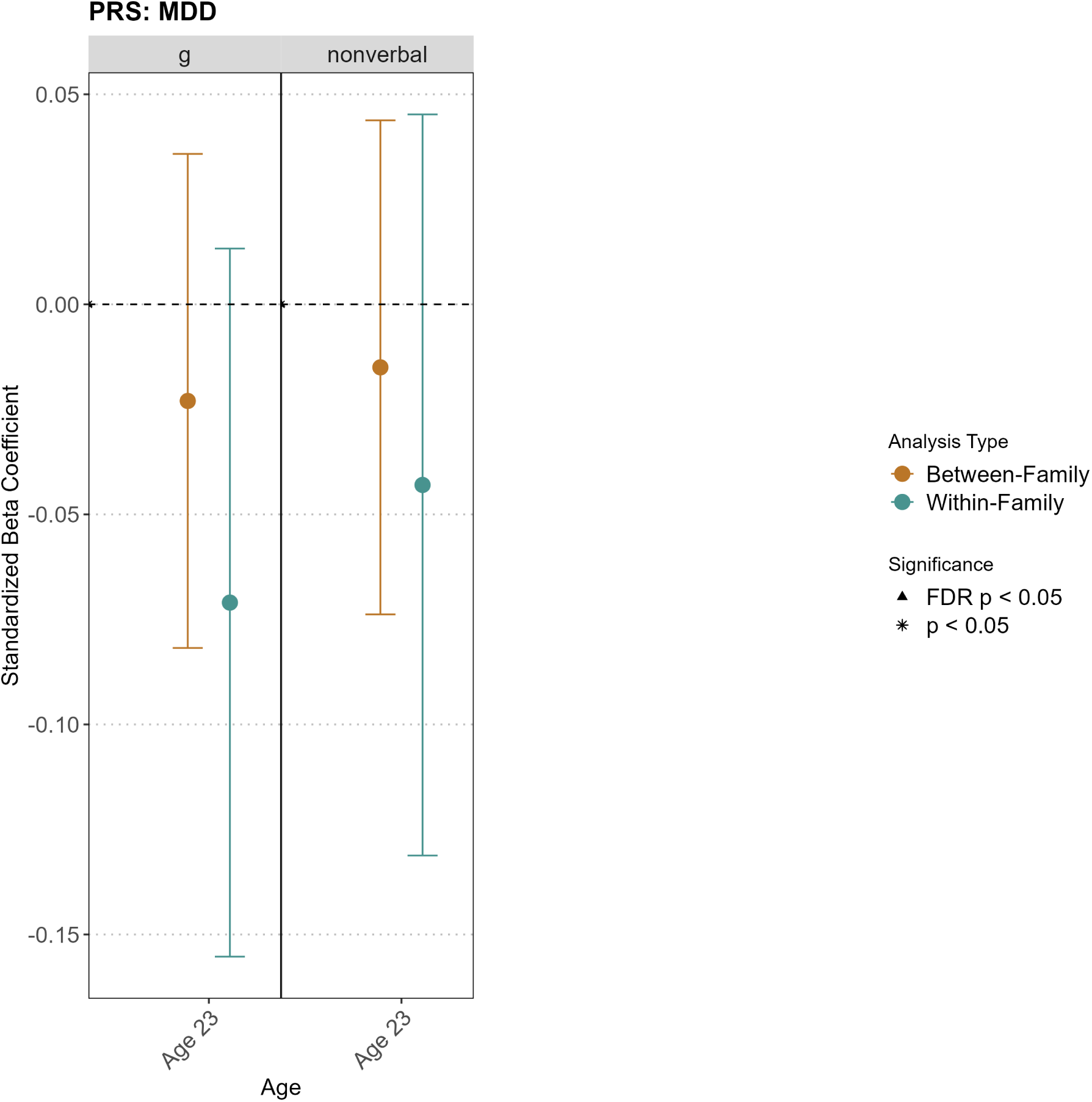
Direct and Indirect Genetic Effects of genetic risk of MDD on Cognitive Abilities.

The figure presents the direct genetic effects and family-mediated indirect genetic effects of major depressive disorder polygenic scores on cognitive abilities across development.

**Figure S12:**
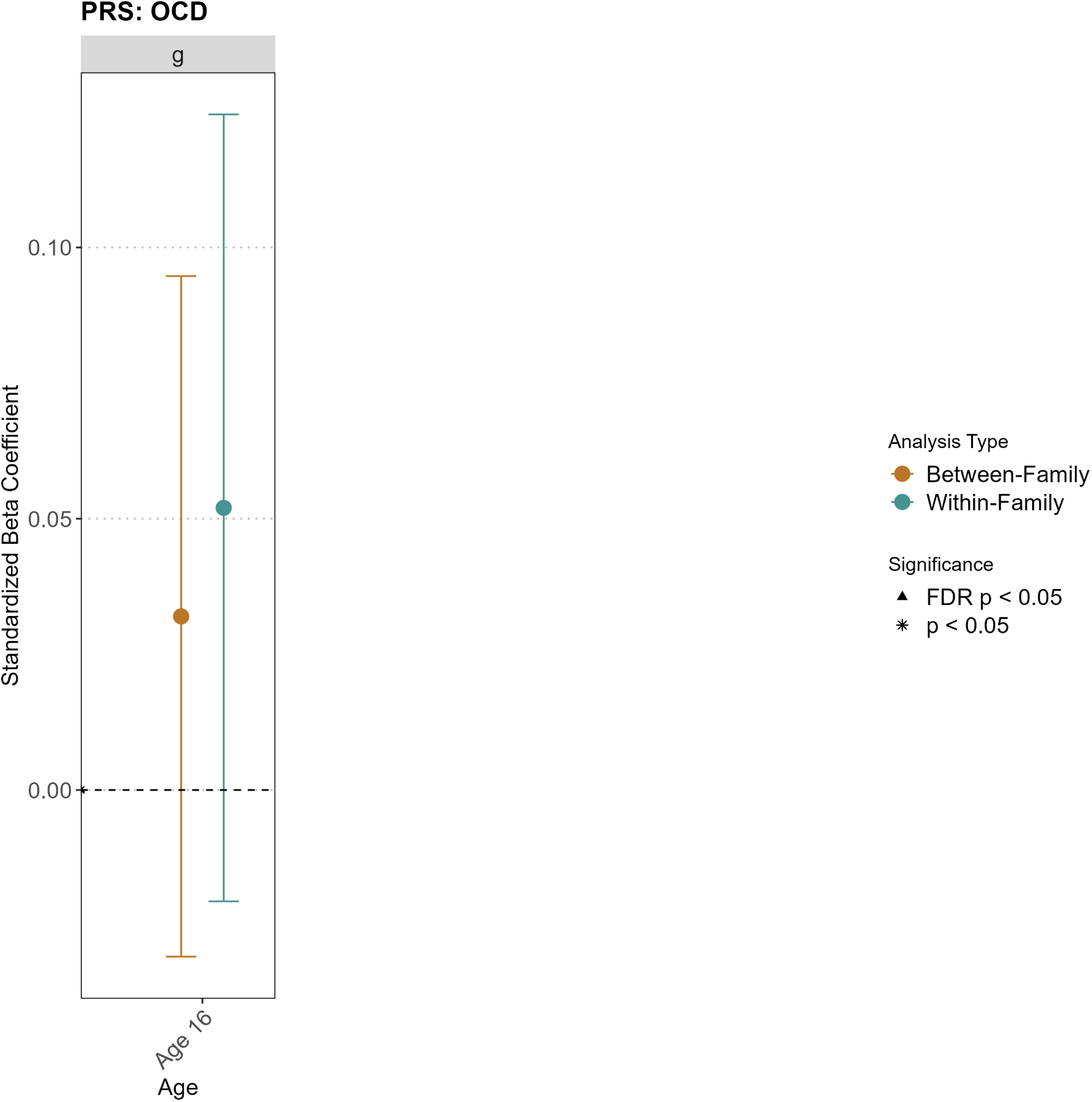
Direct and Indirect Genetic Effects of genetic risk of OCD on Cognitive Abilities.

The figure presents the direct genetic effects and family-mediated indirect genetic effects of obsessive-compulsive disorder polygenic scores on cognitive abilities across development.

**Figure S13:**
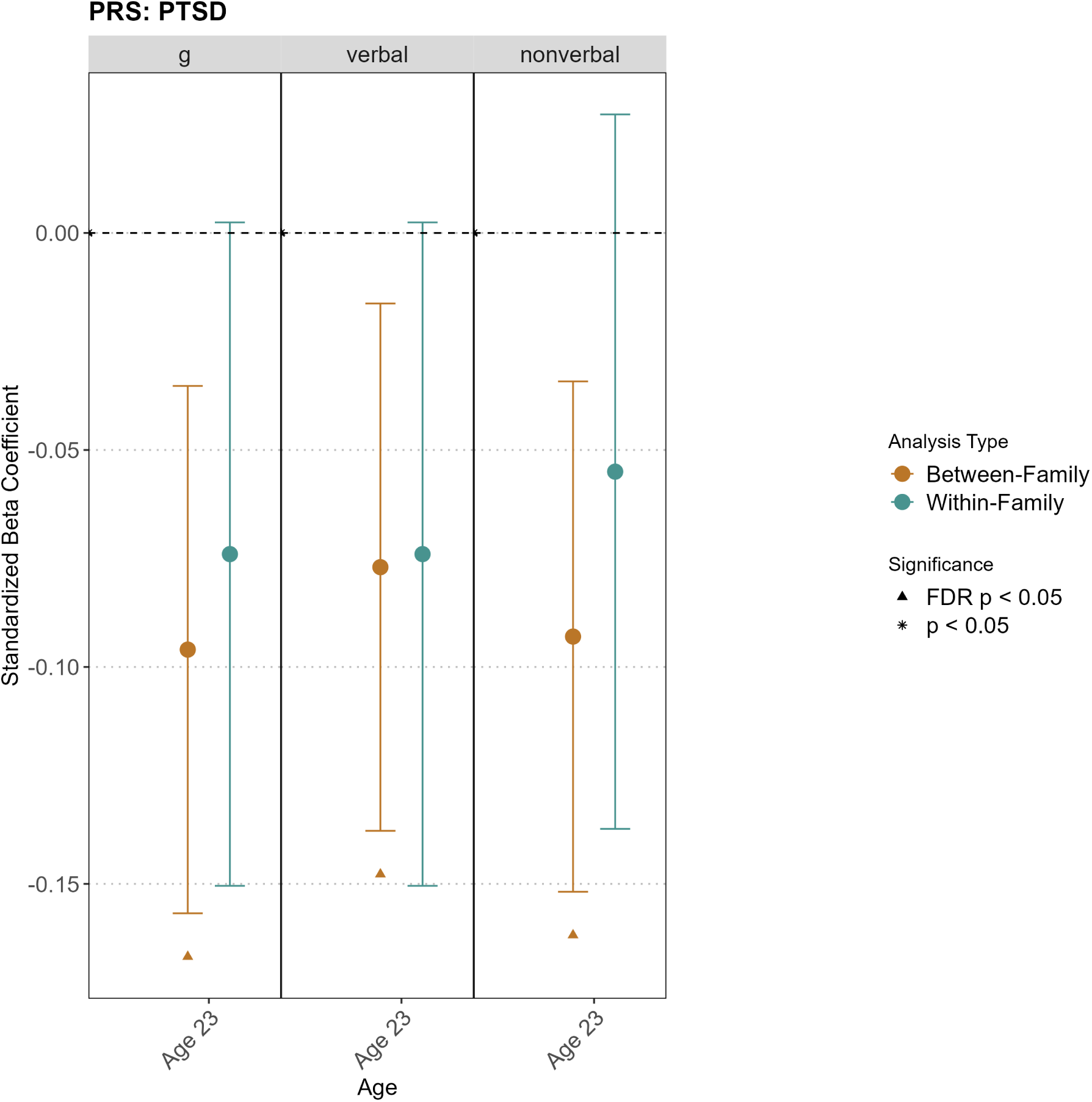
Direct and Indirect Genetic Effects of genetic risk of PTSD on Cognitive Abilities.

The figure presents the direct genetic effects and family-mediated indirect genetic effects of post-traumatic stress disorder polygenic scores on cognitive abilities across development.

**Figure S14:**
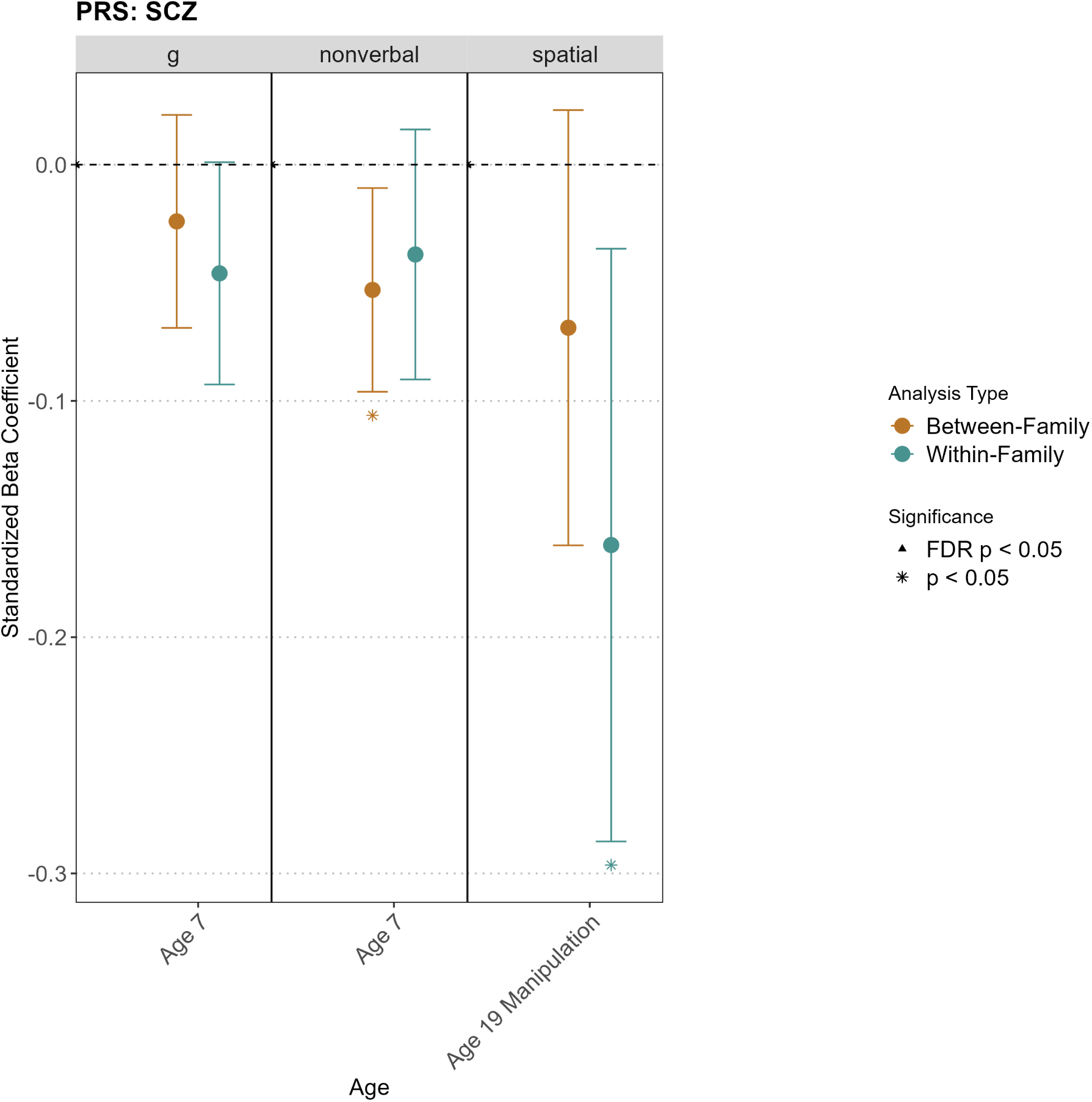
Direct and Indirect Genetic Effects of genetic risk of SCZ on Cognitive Abilities.

The figure presents the direct genetic effects and family-mediated indirect genetic effects of schizophrenia polygenic scores on cognitive abilities across development.

**Figure S15:**
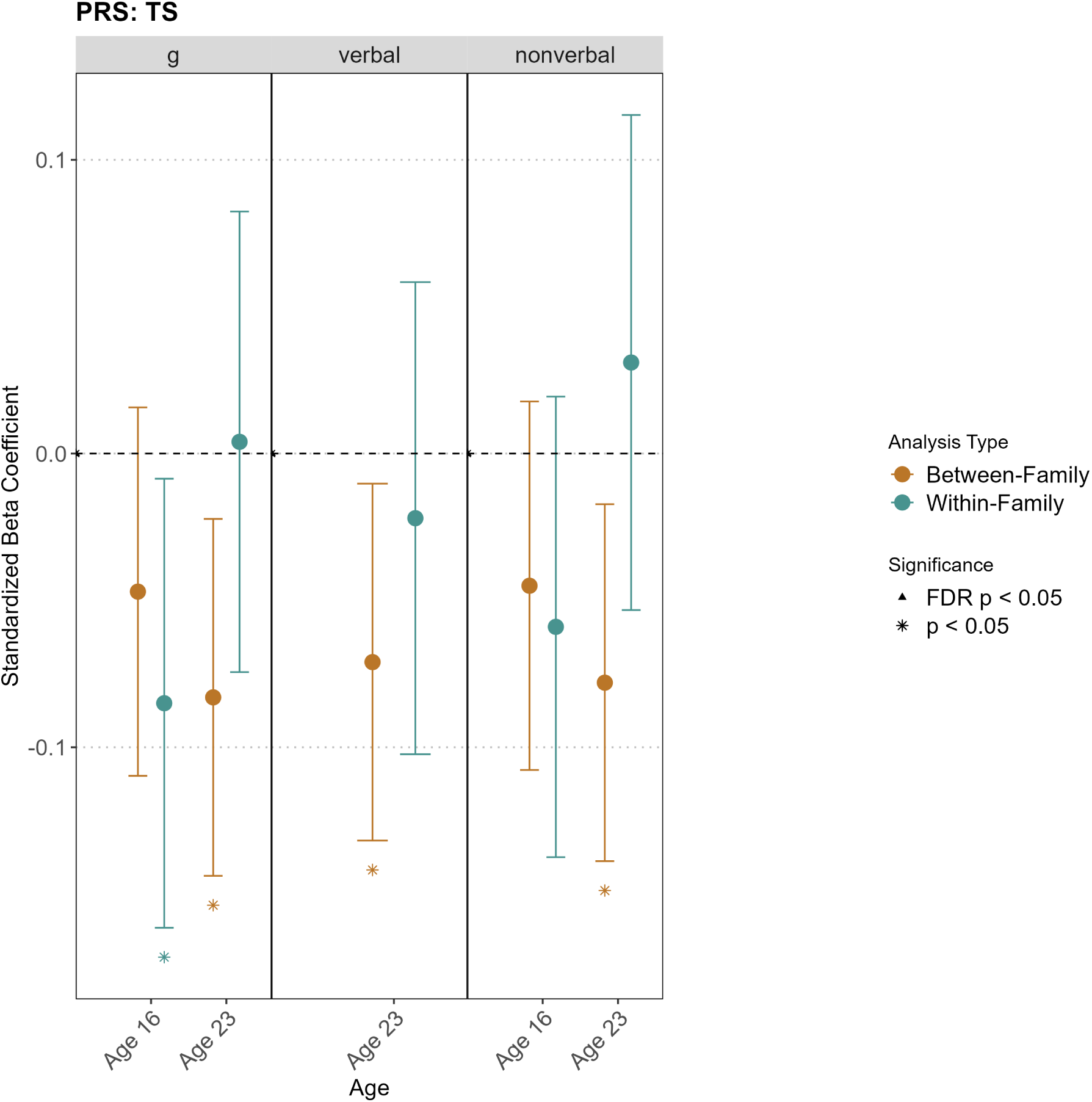
Direct and Indirect Genetic Effects of genetic risk of TS on Cognitive Abilities.

The figure presents the direct genetic effects and family-mediated indirect genetic effects of Tourette syndrome polygenic scores on cognitive abilities across development.

**Figure S16:**
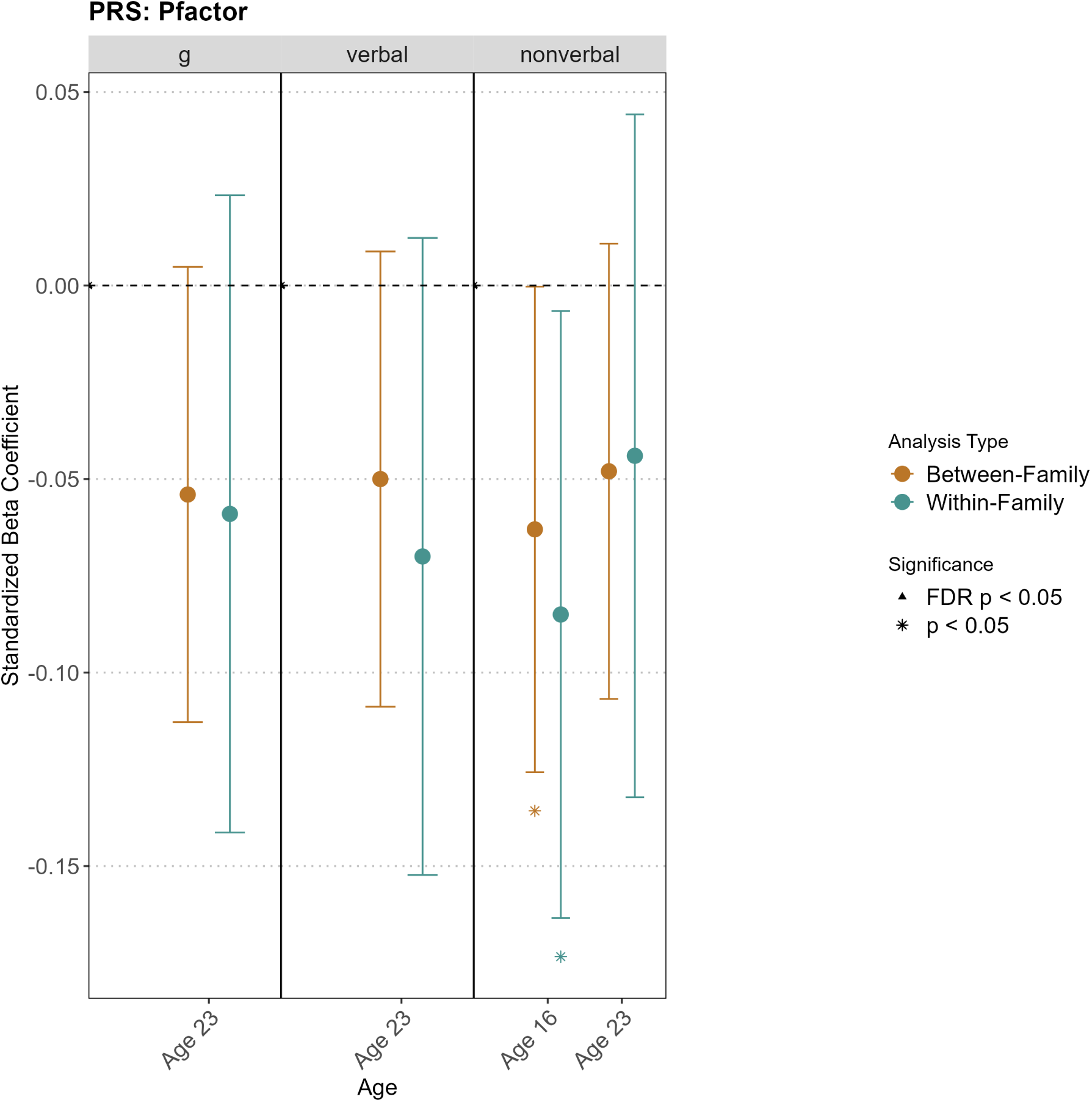
Direct and Indirect Genetic Effects of genetic risk of p factor on Cognitive Abilities.

The figure presents the direct genetic effects and family-mediated indirect genetic effects of transdiagnostic p factor polygenic scores on cognitive abilities across development.

**Figure S17:**
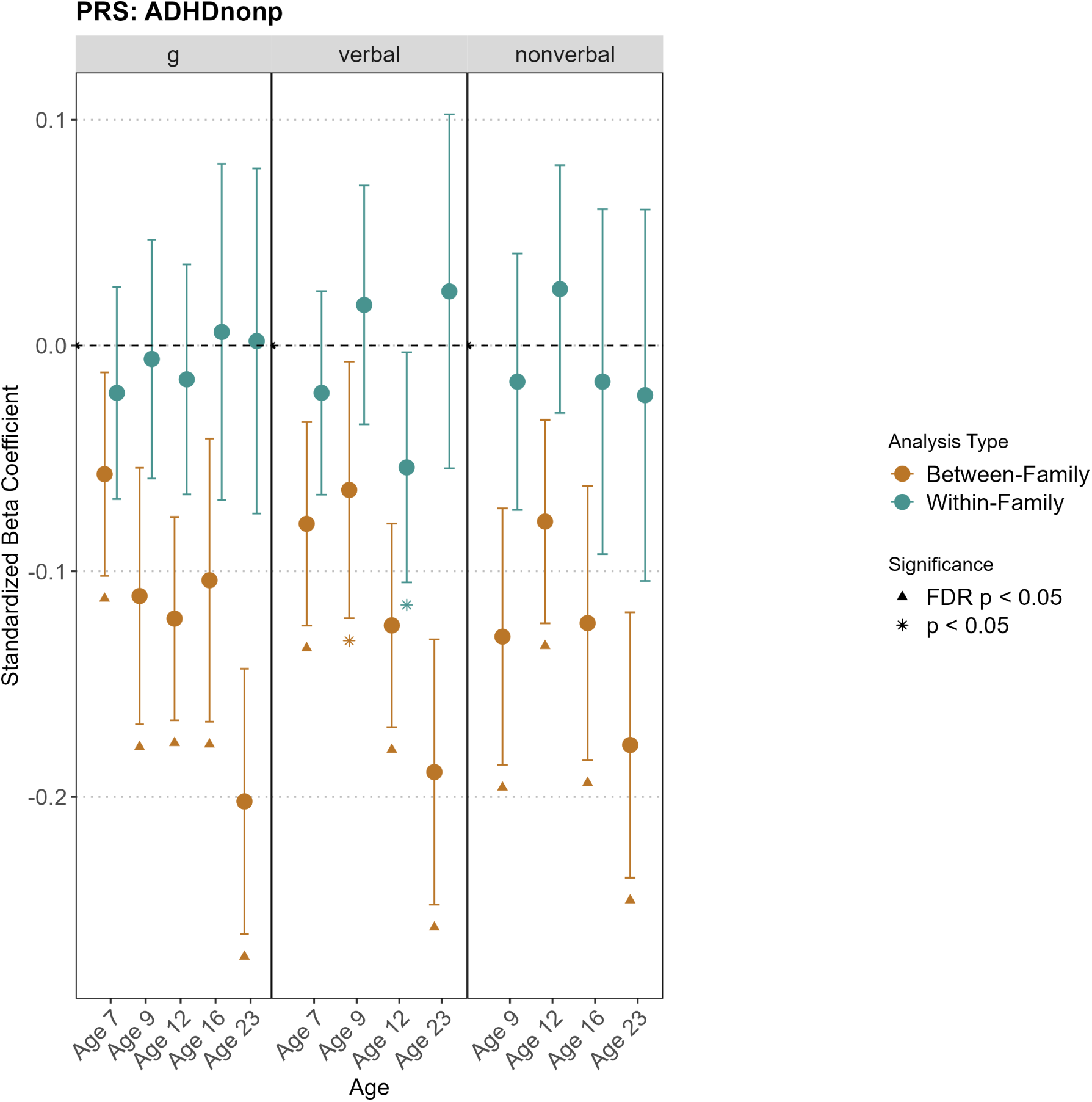
Direct and Indirect Genetic Effects of genetic risk of ADHD non-p on Cognitive Abilities.

The figure presents the direct genetic effects and family-mediated indirect genetic effects of ADHD polygenic scores (corrected for p factor) on cognitive abilities across development.

**Figure S18:**
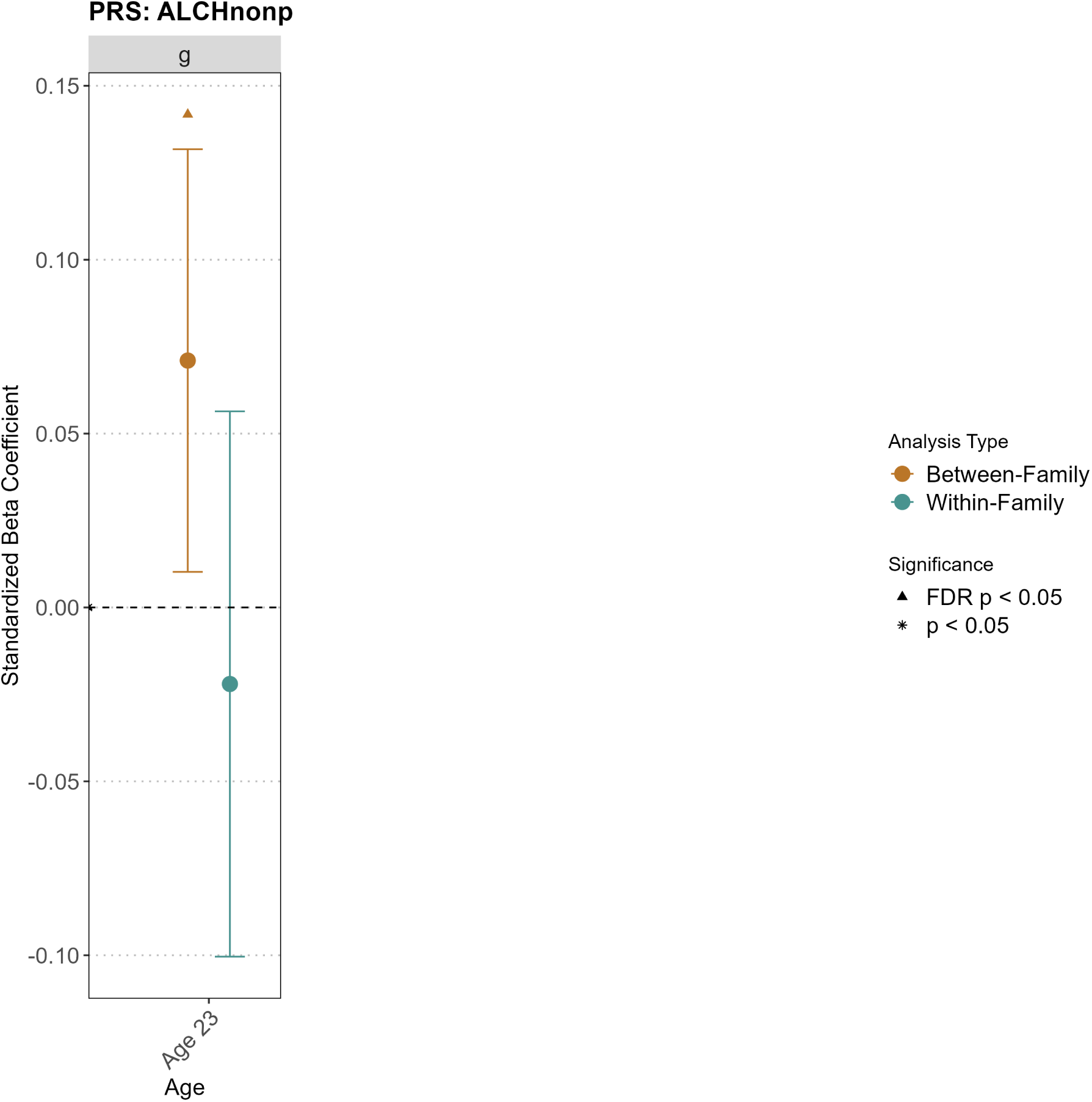
Direct and Indirect Genetic Effects of genetic risk of ALCH non-p on Cognitive Abilities.

The figure presents the direct genetic effects and family-mediated indirect genetic effects of alcohol use disorder polygenic scores (corrected for p factor) on cognitive abilities across development.

**Figure S19:**
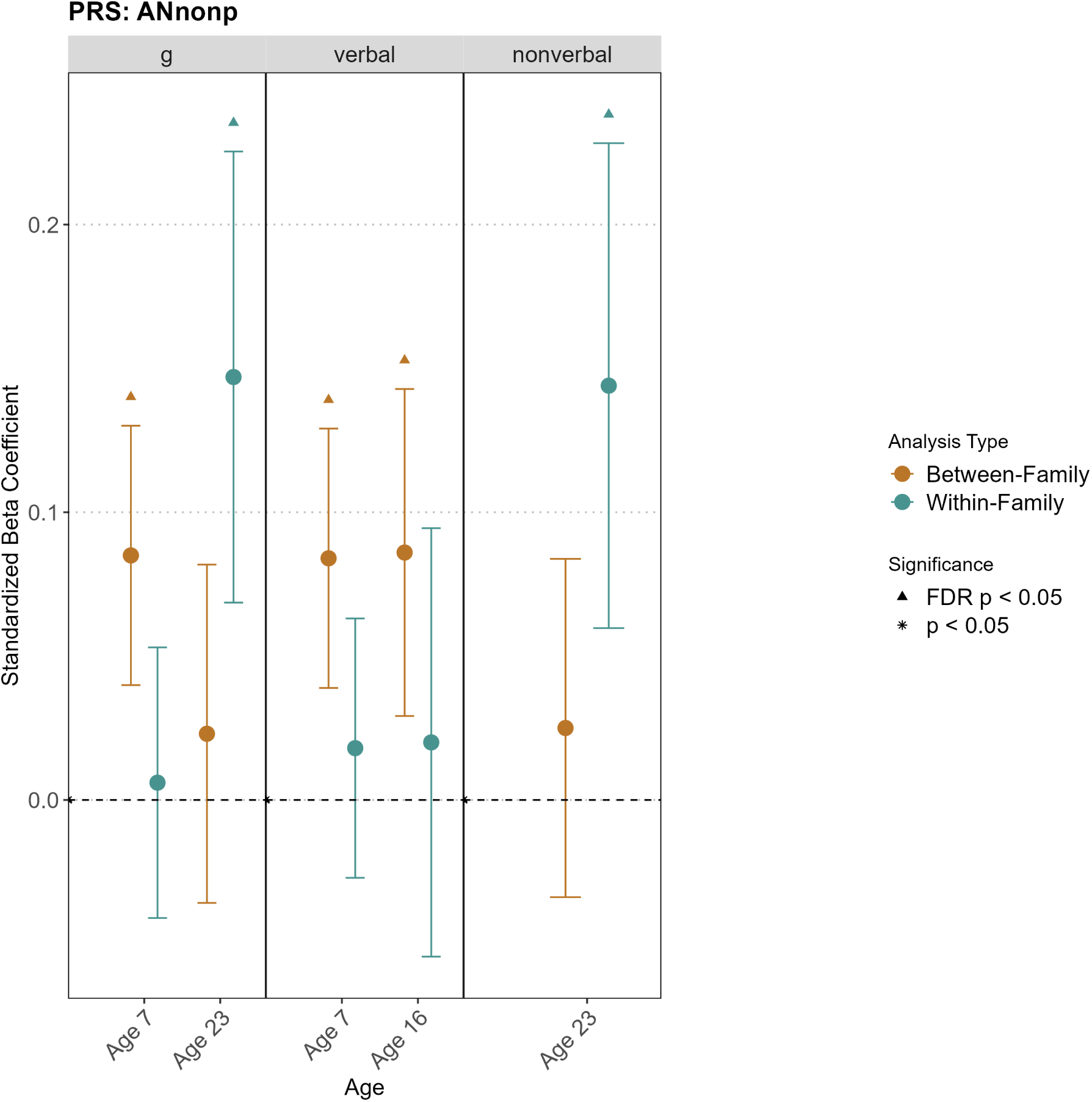
Direct and Indirect Genetic Effects of genetic risk of AN non-p on Cognitive Abilities.

The figure presents the direct genetic effects and family-mediated indirect genetic effects of anorexia nervosa polygenic scores (corrected for p factor) on cognitive abilities across development.

**Figure S20:**
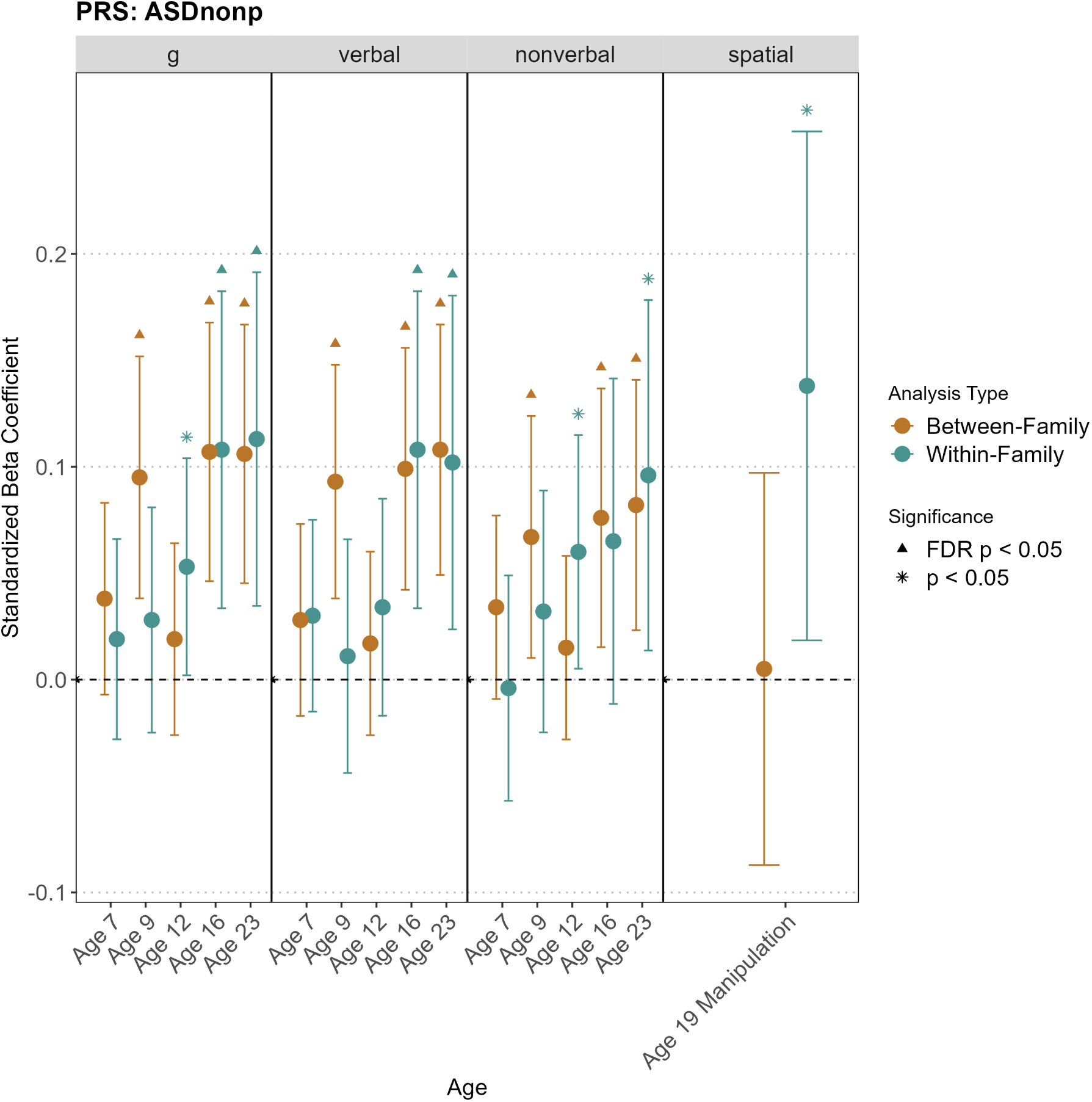
Direct and Indirect Genetic Effects of genetic risk of ASD non-p on Cognitive Abilities.

The figure presents the direct genetic effects and family-mediated indirect genetic effects of autism spectrum disorder polygenic scores (corrected for p factor) on cognitive abilities across development.

**Figure S21:**
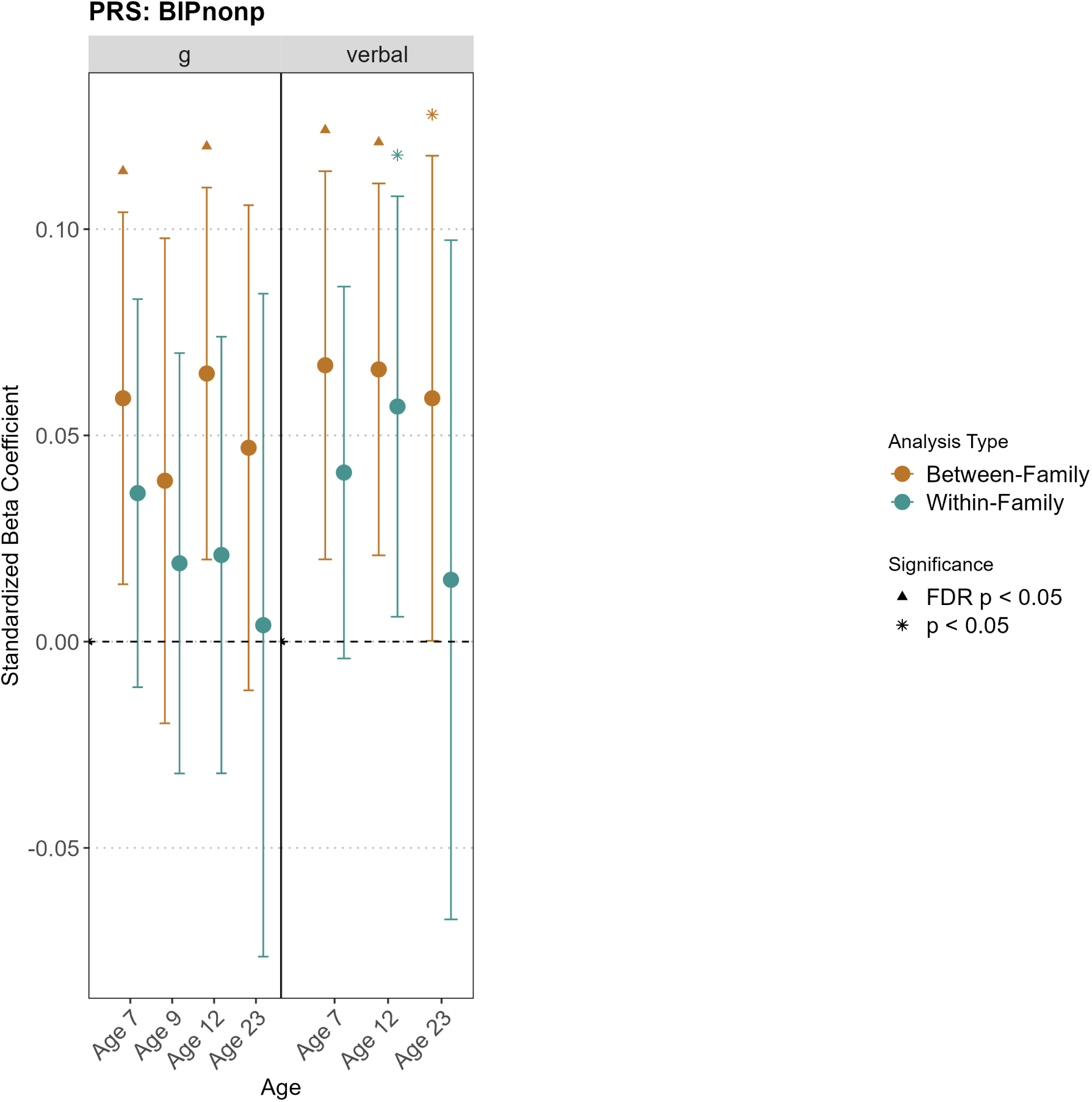
Direct and Indirect Genetic Effects of genetic risk of BIP non-p on Cognitive Abilities.

The figure presents the direct genetic effects and family-mediated indirect genetic effects of bipolar disorder polygenic scores (corrected for p factor) on cognitive abilities across development.

**Figure S22:**
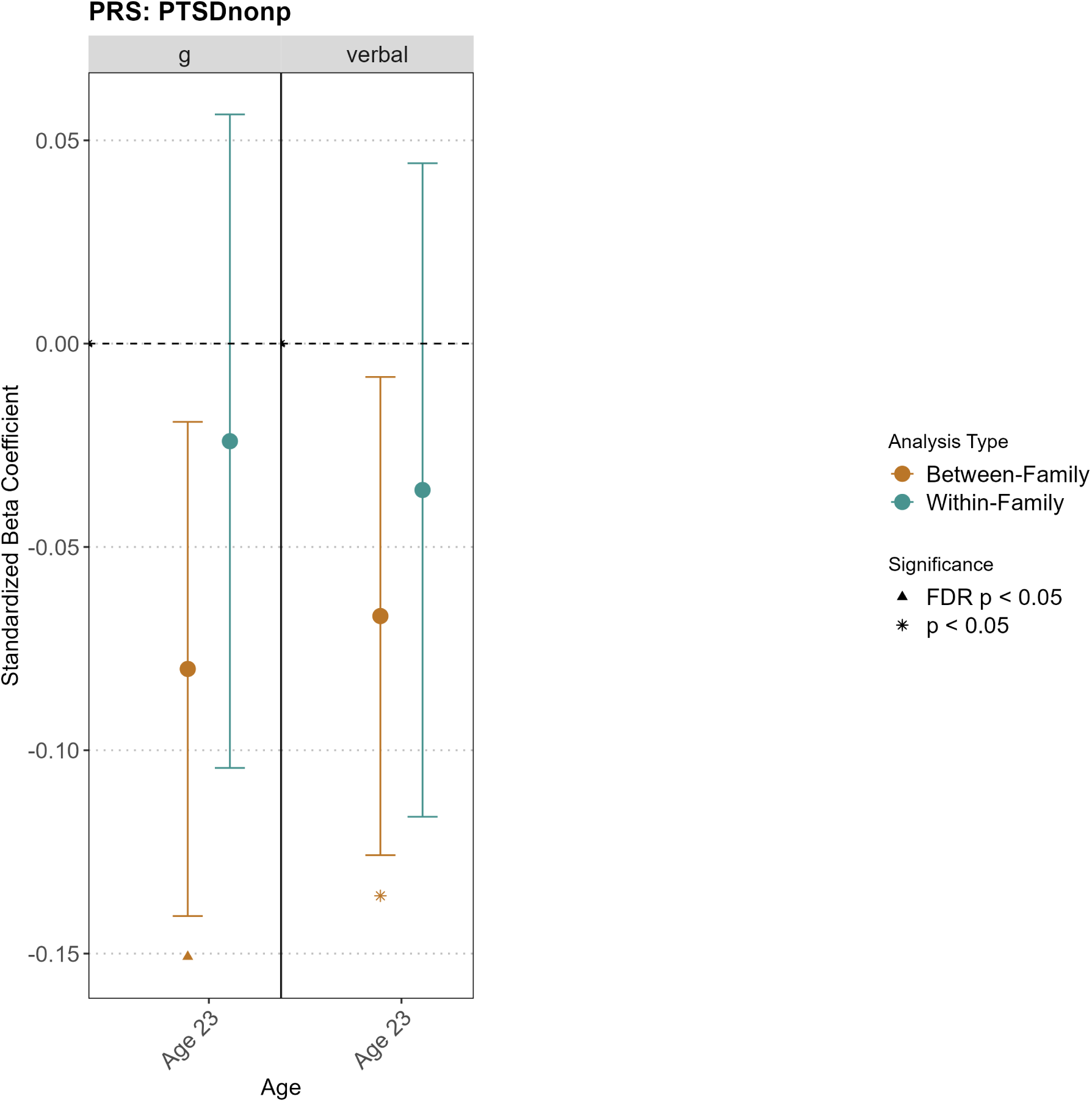
Direct and Indirect Genetic Effects of genetic risk of PTSD non-p on Cognitive Abilities.

The figure presents the direct genetic effects and family-mediated indirect genetic effects of post-traumatic stress disorder polygenic scores (corrected for p factor) on cognitive abilities across development.

**Figure S23:**
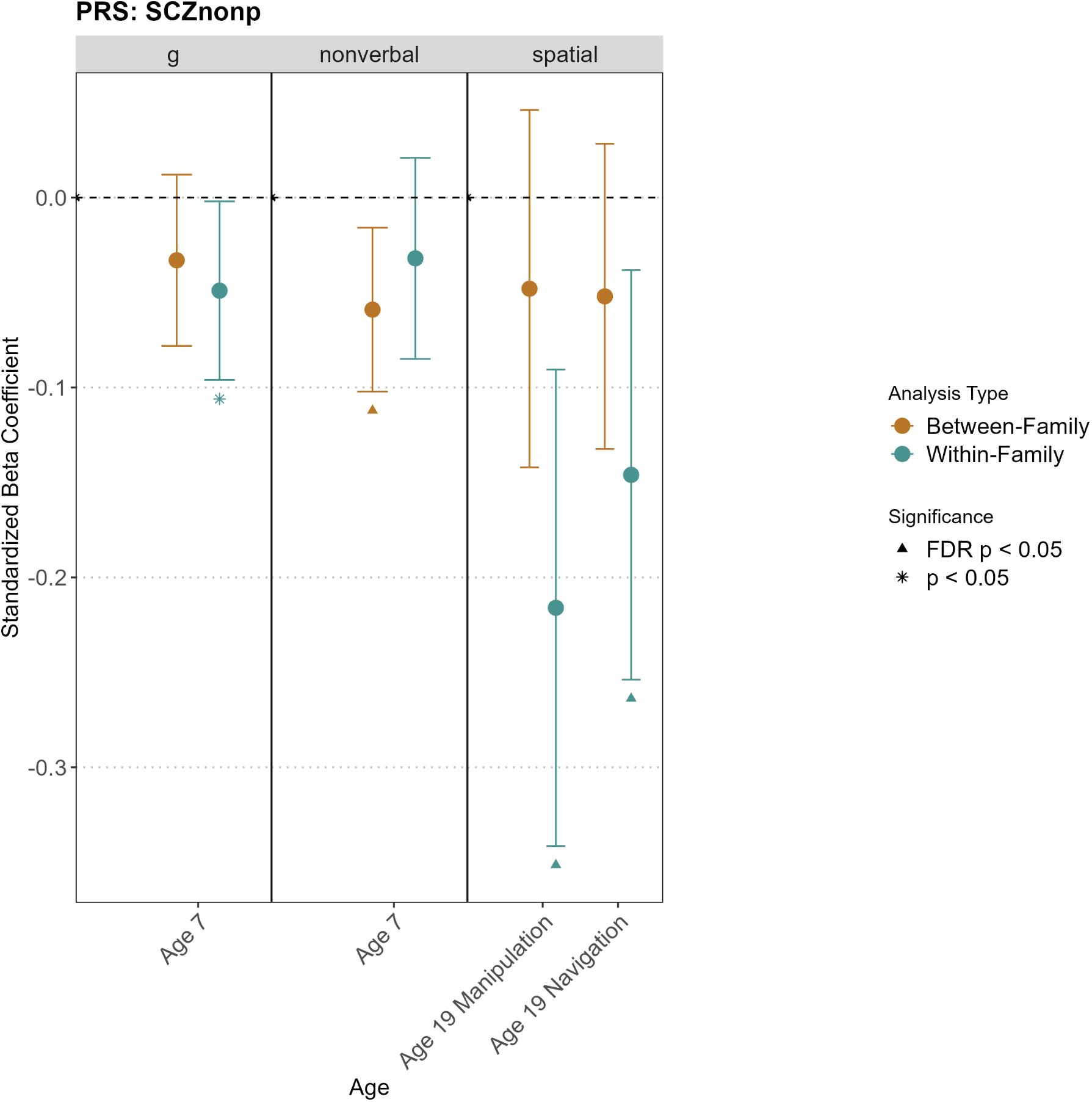
Direct and Indirect Genetic Effects of genetic risk of SCZ non-p on Cognitive Abilities.

The figure presents the direct genetic effects and family-mediated indirect genetic effects of schizophrenia polygenic scores (corrected for p factor) on cognitive abilities across development.

**Figure S24:**
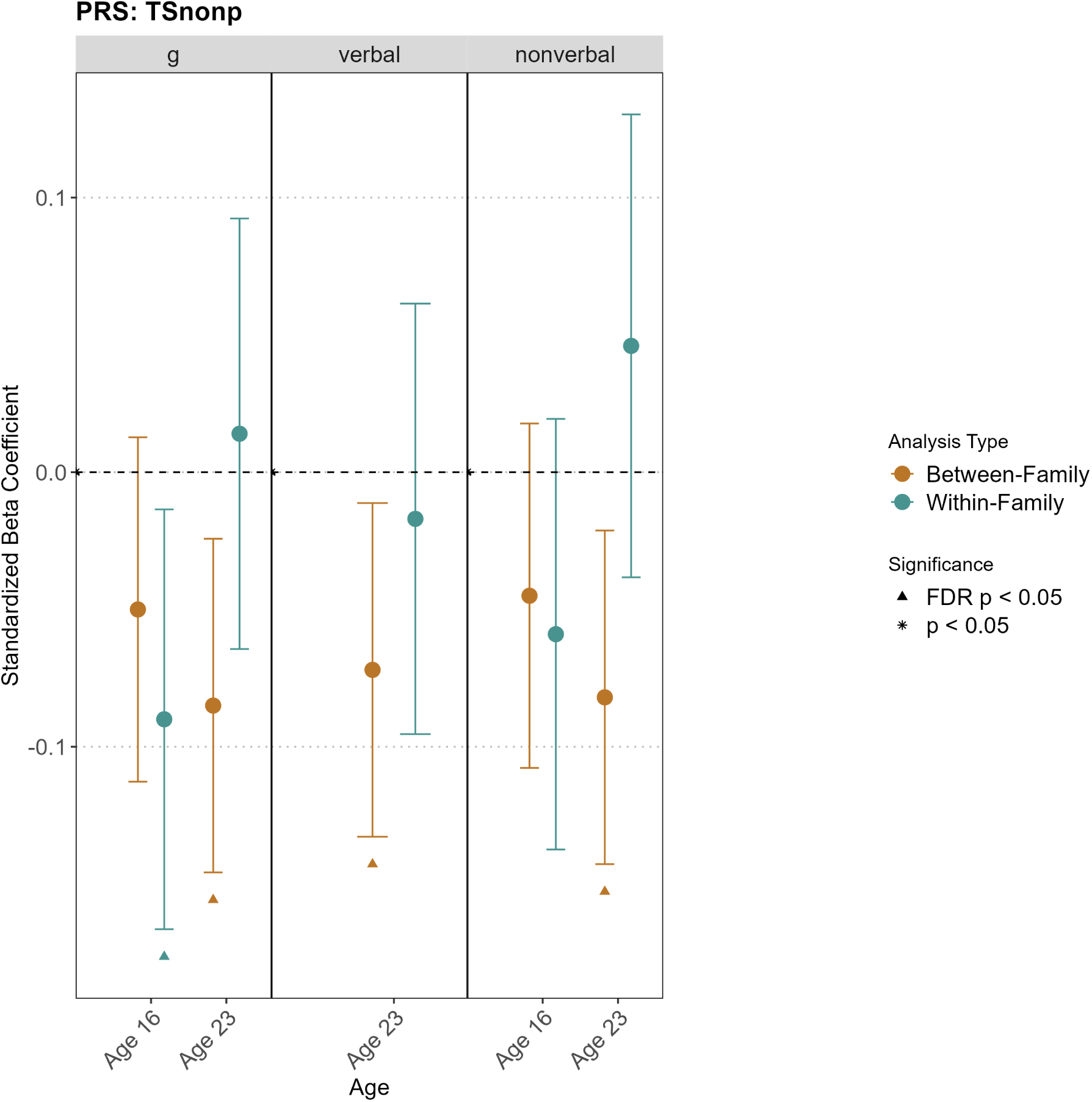
Direct and Indirect Genetic Effects of genetic risk of TS non-p on Cognitive Abilities.

The figure presents the direct genetic effects and family-mediated indirect genetic effects of Tourette syndrome polygenic scores (corrected for p factor) on cognitive abilities across development.

